# A Patient-specific Computational Model for Neonates and Infants with Borderline Left Ventricles

**DOI:** 10.1101/2025.07.15.25331596

**Authors:** Yurui Chen, Isao A. Anzai, David M. Kalfa, Vijay Vedula

## Abstract

**Purpose:** Borderline left ventricle (BLV) presents a dilemma between pursuing a biventricular repair (BiVR) and a Stage 1 palliation (S1P) because a discordant pursuit of BiVR increases mortality risk. We aim to develop and validate a personalized computational model to assist surgical decision-making by predicting virtual surgery hemodynamics in BLV patients.

**Methods:** We developed a novel multi-block lumped parameter network (LPN) model of a BLV circulatory system. Patient-specific model parameters were estimated using a semi-automatic tuning framework to fit clinical data in ten retrospectively identified BLV patients. Virtual surgeries (BiVR and S1P) were performed on each patient to quantify post-operative hemodynamics.

**Results:** In patients who clinically received S1P (Group I, N=5), a virtual BiVR predicted significantly elevated mean pulmonary artery pressure (PAP_mean_: 38.00±10.0 vs. 17.50±2.7 mmHg, *p*<0.01), mean left atrial pressure (LAP_mean_: 25.40±8.2 vs. 6.20±1.2 mmHg, *p*<0.0001), and single ventricle end-diastolic pressure (SVEDP: 21.80±8.7 vs. 4.80±1.3 mmHg, *p*<0.0001) compared with a virtual S1P. A virtual BiVR in patients who clinically underwent BiVR (Group II, N=5) did not predict any adverse hemodynamic outcome.

**Conclusions:** A novel digital twinning framework was developed to predict hemodynamics following virtual surgeries in BLV patients. The model predictions align with the clinically adopted procedure in this retrospectively selected cohort by predicting unacceptable PAP, LAP, and SVEDP. This predictive tool may guide surgeons in determining the hemodynamically optimal surgery for BLV infants, but it needs prospective validation.

**CENTRAL MESSAGE:** Patient-specific computational modeling can predict hemodynamics following virtual surgery in borderline left ventricles and may assist surgical decision-making.

**PERSPECTIVE:** A critical dilemma pediatric heart surgeons and pediatric cardiologists face is choosing between biventricular repair and single ventricle palliation in patients born with a borderline left ventricle. Computational modeling using lumped parameter networks predicts hemodynamics from virtual surgery simulations and may enable clinicians to decide on the hemodynamically optimal procedure.

## INTRODUCTION

Neonates born with underdeveloped left-sided heart structures but not severe enough to be classified as hypoplastic left heart syndrome (HLHS) are categorized as borderline left ventricles (BLV)^1^. BLV patients may manifest variable degrees of hypoplasia of the mitral valve, left ventricle (LV), aortic valve, and aortic arch^1,2^. Suppose the anatomy is adequate and a biventricular repair (BiVR) is deemed feasible without exposing the patient to left atrial hypertension (LAH) while allowing an acceptable cardiac output, a BiVR brings all the benefits of a two-ventricle physiology. In such a situation, a BiVR avoids the multiple surgeries and complications related to single ventricle physiology due to central venous hypertension^3,4^, and lowers the risk of liver disease^5^. Indeed, long-term outcomes for patients undergoing single ventricle palliation remain unsatisfactory, with only 70% reaching adulthood and a high risk of inter-stage morbidity and mortality^6^, and a likelihood of heart or heart/liver failure and transplantation^7,8^.

On the other hand, opting for a BiVR in a BLV neonate who is not a good candidate for a two-ventricle physiology exposes the patient to an immediate high risk of morbidity and mortality related to LAH and low cardiac output syndrome. A conversion from a failing BiVR to a Stage 1 Palliation (S1P) or Norwood operation is also associated with a high risk of mortality^9^. BLV patients undergoing an S1P may also be candidates for a later BiVR following one or multiple staged LV recruitment procedures^10–14^, which also carry a burden of complications^15,16^. The initial decision-making between BiVR and S1P in this patient group is thus critical and conditions both short-term and long-term outcomes.

This initial decision-making for BLV patients is not only critical but also remains controversial and very difficult. The current surgical decision-making primarily relies on *status quo* echo-based or MRI-based morphological and functional measurements of the heart and related vasculature, and an overall subjective approach to the anatomy and physiology of each patient^17–22^. Various echo-based indices have been developed, but have limited clinical utility or use by the medical and surgical community^23–27^. A comprehensive BSA-weighted score was developed based on the mitral valve area, LV long-axis dimensions relative to the heart, and the diameter of the aortic root to determine the surgery to be performed^27^. However, scores based on the chamber volume measurements using the classical Simpson’s method may have limited applicability for BLV patients because of the differences in the shape of the hypoplastic LV from that of the normal LV^26^. Moreover, these morphology-based scores and indices do not take into account the specific anatomy/defects and physiology of each patient in a personalized way. Further, providing pediatric cardiac surgeons and cardiologists the ability to include predicted post-surgical hemodynamics, such as the pressures in the left atrium (LA), pulmonary artery (PA), and the systemic ventricle and ventricular stroke volume, as part of the decision-making could transform the treatment planning and care for BLV patients.

Prior studies have applied blood flow modeling to various stages of single ventricle palliation in patients with HLHS (the most severe end of the BLV spectrum), including Stage 1 (Norwood)^28,29^, Stage II (Glenn) ^30,31^, and Stage III (Fontan) procedures^32–34^. However, modeling the circulation of BLV patients (not HLHS) has not received much attention. In this study, we aim to (a) develop a novel patient-specific lumped parameter network (LPN) model of the circulatory system for BLV patients, (b) perform virtual BiVR and S1P surgeries using this model in ten BLV patients identified retrospectively, and (c) assess whether the model-predicted post-operative hemodynamics corroborate retrospectively the decision made by the multi-disciplinary clinical team by showing unacceptably high LA and PA pressures after virtual BiVR in patients who were clinically offered an S1P. We will also validate our model-predicted virtual surgery hemodynamics in a subcohort of patients for whom post-operatively acquired clinical data is available.

## METHODS

### Study Population

We included retrospectively the ten most recent infants diagnosed with BLV (defined as mitral valve annulus Z-score between -2 and -5 and/or aortic annulus Z-score between -2 and -5 and/or indexed left ventricle end-diastolic volume (LVEDVi) Z-score between -2 and -5 on transthoracic echocardiogram) who underwent either a BiVR or an S1P procedure at the Columbia University Irving Medical Center (CUIMC). Patients with measurements below Z-score -5 or above -2 were omitted. This single-center study was approved by the CUIMC Institutional Review Board (IRB# AAAT2664 approved on July 10, 2020), while obtaining informed consent was waived due to the study being retrospective. Five patients (Group I, age 2.0±0.6 months) underwent the S1P procedure, and the remaining five (Group II, age 1.6±1.2 months) experienced BiVR. The patients’ clinical, echocardiographic, and hemodynamic characteristics are summarized in **Table 1**, and the raw data for all the patients in the study cohort is shared in **Supplementary Table ST1**.

**Table 1.**
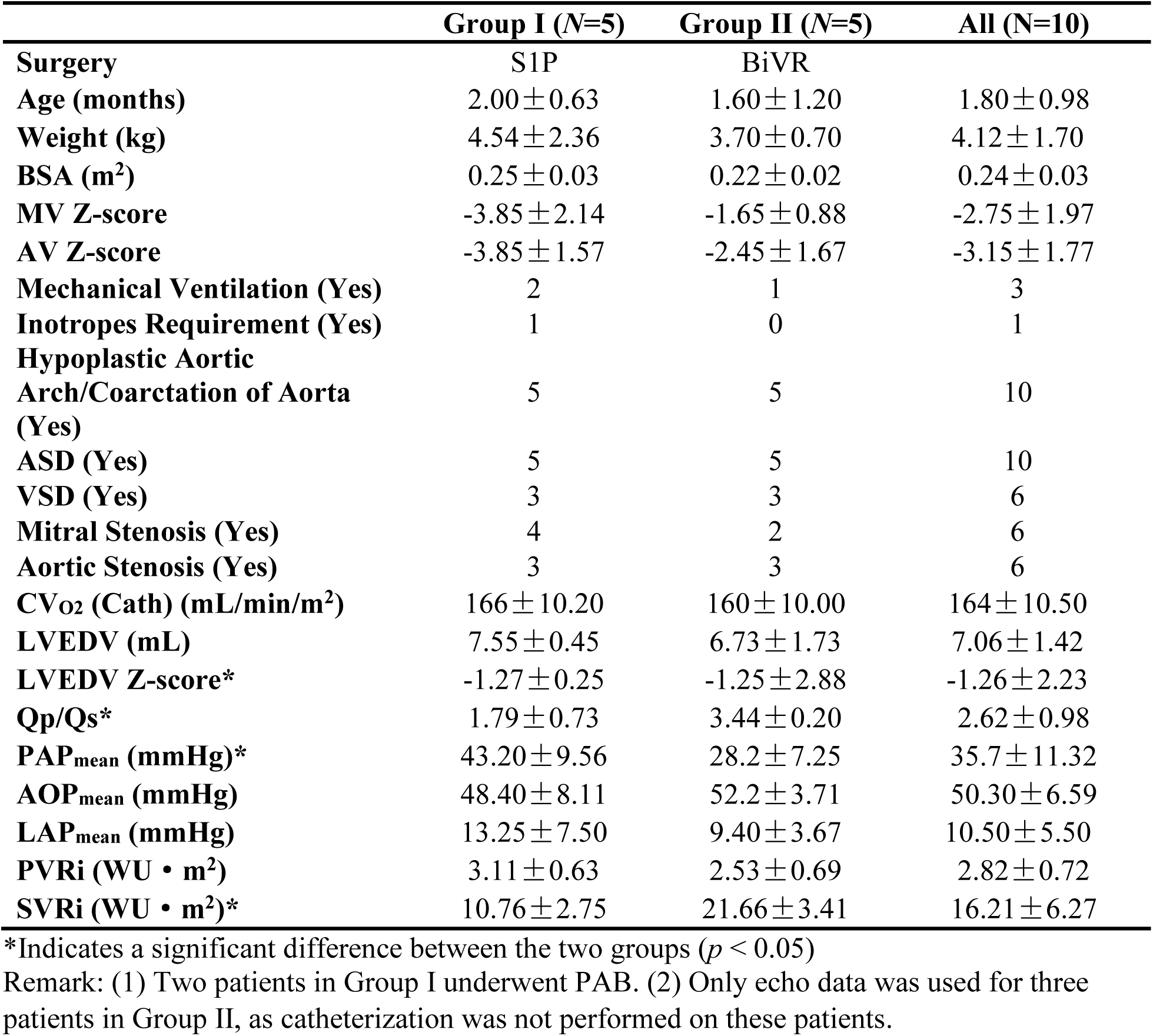
Preoperative clinical characteristics for the BLV patients included in this study.

|  | **Group I (N=5)** | **Group II (N=5)** | **All (N=10)** |
| --- | --- | --- | --- |
| **Surgery** | S1P | BiVR |  |
| **Age (months)** | 2.00 ± 0.63 | 1.60 ± 1.20 | 1.80 ± 0.98 |
| **Weight (kg)** | 4.54 ± 2.36 | 3.70 ± 0.70 | 4.12 ± 1.70 |
| **BSA (m<sup>2</sup>)** | 0.25 ± 0.03 | 0.22 ± 0.02 | 0.24 ± 0.03 |
| **MV Z-score** | -3.85 ± 2.14 | -1.65 ± 0.88 | -2.75 ± 1.97 |
| **AV Z-score** | -3.85 ± 1.57 | -2.45 ± 1.67 | -3.15 ± 1.77 |
| **Mechanical Ventilation (Yes)** | 2 | 1 | 3 |
| **Inotropes Requirement (Yes)** | 1 | 0 | 1 |
| **Hypoplastic Aortic Arch/Coarctation of Aorta (Yes)** | 5 | 5 | 10 |
| **ASD (Yes)** | 5 | 5 | 10 |
| **VSD (Yes)** | 3 | 3 | 6 |
| **Mitral Stenosis (Yes)** | 4 | 2 | 6 |
| **Aortic Stenosis (Yes)** | 3 | 3 | 6 |
| **CV<sub>O2</sub> (Cath) (mL/min/m<sup>2</sup>)** | 166 ± 10.20 | 160 ± 10.00 | 164 ± 10.50 |
| **LVEDV (mL)** | 7.55 ± 0.45 | 6.73 ± 1.73 | 7.06 ± 1.42 |
| **LVEDV Z-score\*** | -1.27 ± 0.25 | -1.25 ± 2.88 | -1.26 ± 2.23 |
| **Qp/Qs\*** | 1.79 ± 0.73 | 3.44 ± 0.20 | 2.62 ± 0.98 |
| **PAP<sub>mean</sub> (mmHg)\*** | 43.20 ± 9.56 | 28.2 ± 7.25 | 35.7 ± 11.32 |
| **AOP<sub>mean</sub> (mmHg)** | 48.40 ± 8.11 | 52.2 ± 3.71 | 50.30 ± 6.59 |
| **LAP<sub>mean</sub> (mmHg)** | 13.25 ± 7.50 | 9.40 ± 3.67 | 10.50 ± 5.50 |
| **PVRi (WU • m<sup>2</sup>)** | 3.11 ± 0.63 | 2.53 ± 0.69 | 2.82 ± 0.72 |
| **SVRi (WU • m<sup>2</sup>)\*** | 10.76 ± 2.75 | 21.66 ± 3.41 | 16.21 ± 6.27 |

\*Indicates a significant difference between the two groups ( $p < 0.05$ )
Remark: (1) Two patients in Group I underwent PAB. (2) Only echo data was used for three patients in Group II, as catheterization was not performed on these patients.

### Preoperative Circulation Model

We developed an LPN model of the circulatory system for BLV patients, adapted from the model proposed in Corsini et al.^30^ for Norwood (i.e., S1P) circulation, and tailored to each patient’s anatomy and physiology (**Figure 1**). Specific changes were made to match the preoperative BLV patient’s physiology, including (a) reintroducing a heart with two atria and two ventricles modeled using time-varying elastance functions for each chamber; (b) adding resistive elements to mimic shunt flow due to septal defects between the connecting chambers; (c) addition of a resistive element in the ascending aorta proximal to the ventricle to mimic aortic arch hypoplasia; (d) removal of the Blalock-Taussig-Thomas (BTT) shunt with an unclosed patent ductus arteriosus (PDA) between the ascending aorta and pulmonary artery; (e) adding a resistive element to mimic pulmonary artery banding, if applicable; and (f) employing a versatile valve model that simulates the effects of stenosis and regurgitation^35,36^. The LPN model is also coupled to an oxygen transport model that computes arterial and venous saturations (S_art_ and S_ven_) and systemic oxygen delivery (O_2_D)^37^. Details of the LPN model, along with the mathematical expressions and governing differential algebraic equations, are provided in **Supplementary Material Section B**.

**Figure 1.**
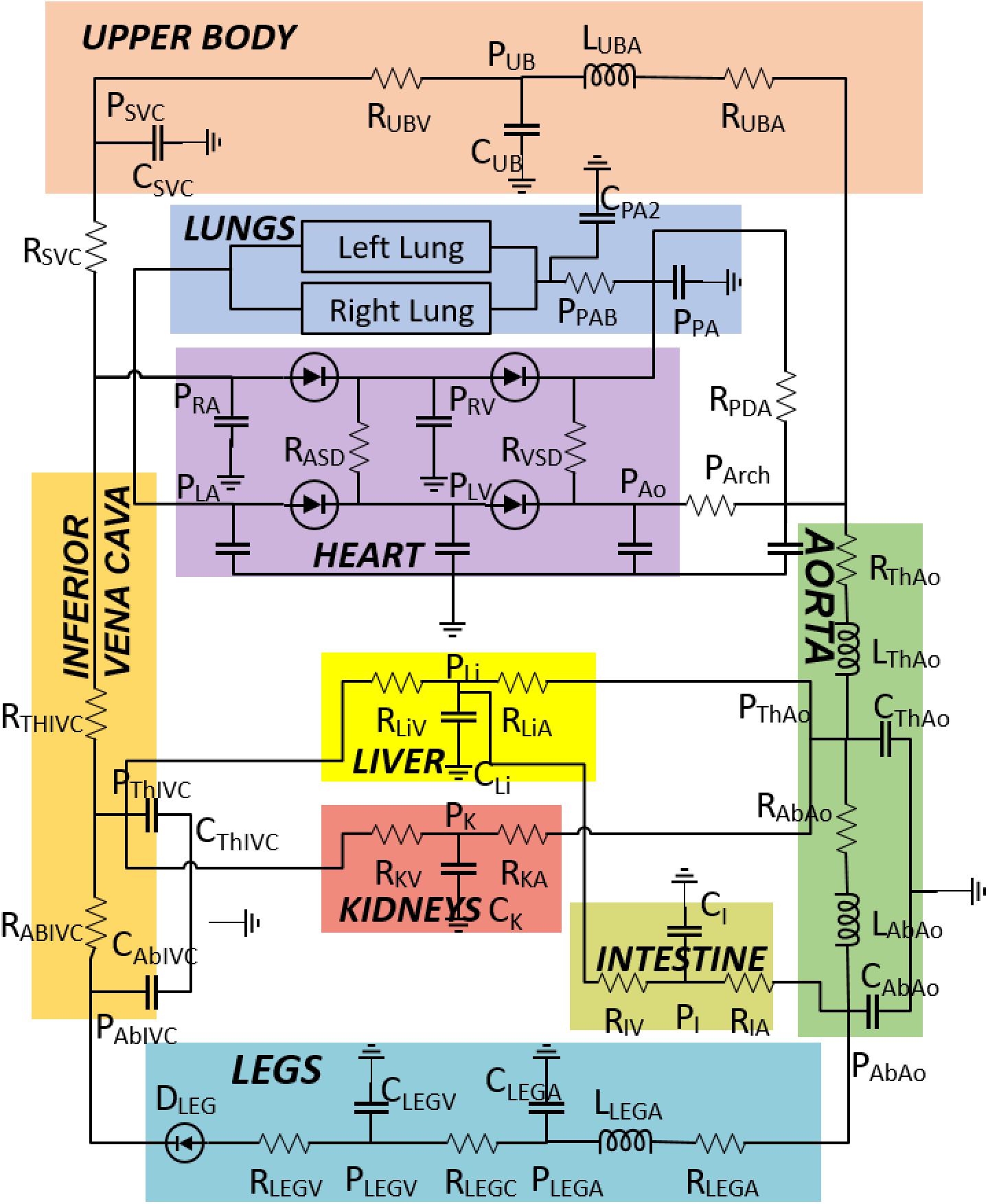
A preoperative multi-block lumped parameter network (LPN) model of the circulatory system for patients with a borderline left ventricle (BLV). *C*: capacitance or vessel wall compliance; *R*: resistance to blood flow; *L*: inductance or fluid inertia; *P*: pressure. **Upper Body:** *SVC*, superior vena cava; *UB*, upper body; *UBA*, upper body arteries; *UBV*, upper body veins; **Lungs:** *PA*, pulmonary artery; *PAB*, pulmonary artery banding (if applicable); **Heart:** *RA*, right atrium; *RV*, right ventricle; *LA*, left atrium; *LV*, left ventricle; *ASD*, atrial septal defect; *VSD*, ventricular septal defect; **Aorta**: *Ao*, proximal ascending aorta; *Arch*, aortic arch; *ThAo*, thoracic aorta; *AbAo*, abdominal aorta; **Lower Body:** *LEGA*, arteries in legs; *LEGV*, veins in legs; *AbIVC*, abdominal inferior vena cava; *ThIVC*, thoracic inferior vena cava; *Li*, liver; *LiA*, hepatic arteries; *LiV*, hepatic veins; *K*, kidneys; *KA*, renal arteries; *KV*, renal veins; *I*, intestine; *IA*, intestine artery; *IV*, intestine veins.

### LPN Parameter Estimation

The LPN parameter estimation aims to ensure that the computational model replicates the patient’s preoperative hemodynamic state, enabling patient-specific assessment and treatment planning. A detailed description of the steps (flowchart in **Figure 2**) involved in estimating LPN parameters that fit the BLV patient’s clinical data is provided below^38^:

1. We extracted the patient’s clinical data, including BSA, valve orifice dimensions, vascular resistances, presence of septal defects, pulmonary arterial banding, and hemodynamic data from echocardiographic and catheterization reports. Specifically, data from the catheterization reports include min/max/mean aortic and pulmonary artery pressures (AOP_min/max/mean_, PAP_min/max/mean_), mean left atrial pressure (LAP_mean_), the pulmonary to systemic flow ratio (Qp/Qs), and the pulmonary and systemic vascular resistances (PVR, SVR). Data from echocardiographic reports include the ventricular stroke volume (SV), ejection fraction (EF), and pressure gradients across cardiac valves, septal defects, and PDA. For the patients who only had echo data, we estimated pressures (AOP, PAP, LAP), Qp/Qs, and vascular resistances (PVR, SVR) using the approach described in **Supplementary Material Section C**. These clinical measurements are set as ‘hemodynamic targets’ for the optimization algorithm (discussed in Step 3) to iteratively vary the LPN model parameters so that the model’s hemodynamic predictions closely fit the clinical data.
2. A two-part scaling approach is then performed to define the LPN model parameters matched to the patient’s BSA and vascular resistances^39^.
  i. First, starting with the LPN parameters of a reference adult, based on the model proposed in Snyder & Rideout^40^, we performed a BSA-based allometric scaling to account for the patient’s age-dependent organ/vessel growth as,

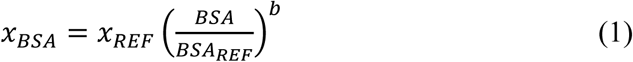 where *x* represents a generic LPN model parameter, such as resistance, capacitance, and inductance, and the subscript REF indicates the corresponding parameter of the reference adult^40^. The exponent *b* depends on the type of the LPN component (i.e., resistance, capacitance, inductance) and the organ/vessel, extracted from Table 2 of Pennati et al.^41^
  ii. Second, we scaled the resistances and capacitances to match the patient’s systemic and pulmonary vascular resistances (SVR, PVR) as,

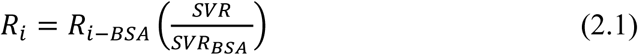

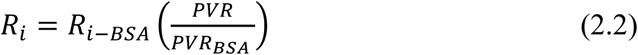

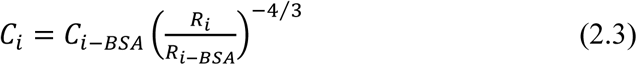 where *R*_*i*_ and *C*_*i*_ indicate the individual resistances and capacitances of the LPN model, while *R*_*i*−*BSA*_ and *C*_*i*−*BSA*_ are the BSA-scaled LPN parameters from the previous step. The exponent (-4/3) was adopted in Eq. (2.3) based on the dependence of the vascular resistance and compliance on the vessel radius and length^30,41–43^
3. At the end of the previous step, we obtained the vascular parameters of the preoperative LPN model. We then used an optimization method to automatically estimate the remaining model parameters, including the heart model and arterial parameters, by minimizing the cost function,

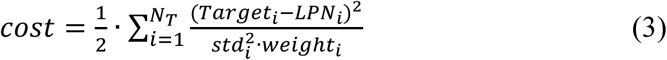 where N_T_ is the total number of hemodynamic targets. The cost function is a weighted sum of squared differences between each clinical target and the corresponding LPN-predicted value. In Eq. (3), *std* and *weight* represent the estimated standard deviation of each clinical measurement and a weighting factor that signifies the clinical importance of that particular quantity^44,45^. These values (*std*, *weight*) were heuristically set based on previously published data^44,45^. The raw data of each patient’s hemodynamic target, along with the corresponding standard deviation and weight factors, is provided in **Supplementary Tables ST2 and ST3**. In this study, we used uniform weights and standard deviations for measurements from a single modality. For example, quantities extracted from cath reports have a higher weight (100%) and lower standard deviation (10%). In comparison, data estimated from echo reports have a lower weight (50%) and higher standard deviation (20%) to account for the noise and measurement uncertainty. Further, we set bounds for each LPN parameter based on previous manual tuning experience to avoid the optimizer producing unphysiological results. We then employ the Nelder-Mead optimization method^46^ to minimize the cost function (Eq. (3)), implemented in an in-house Python-based code, thereby leading to an optimal set of parameters that resulted in a reasonable agreement between model predictions and clinical measurements.

**Figure 2.**
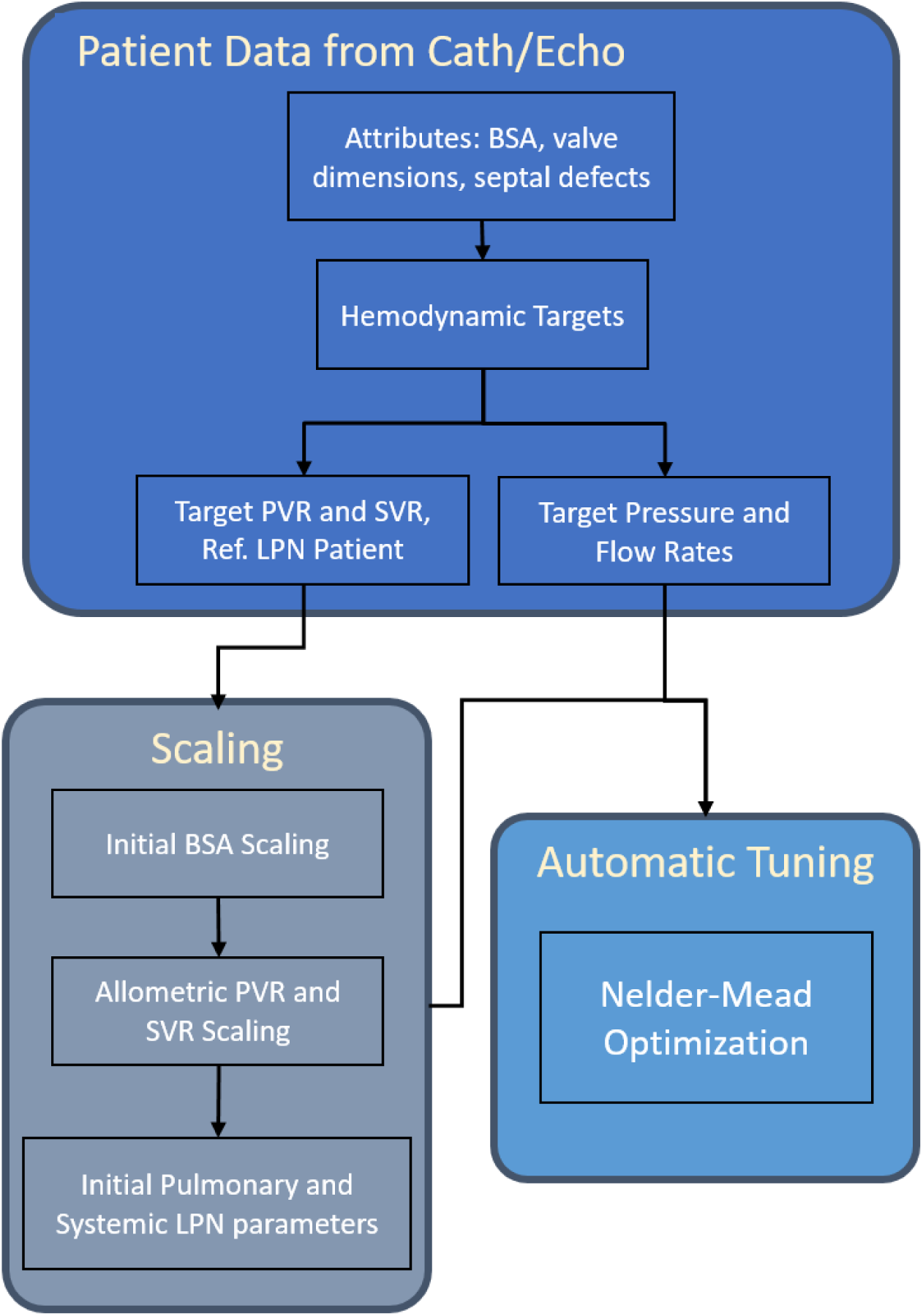
A flowchart of the steps involved in the semi-automatic tuning framework for estimating LPN model parameters. LPN: lumped parameter network; BSA: body surface area; PVR: pulmonary vascular resistance; SVR: systemic vascular resistance.

**Table 2.** Preoperative clinical measurements are set as hemodynamic targets for the LPN model parameter estimation algorithm. The raw data for each patient-specific target value is provided in Supplemental Material.

| Target | Description | Weight | STD |
| --- | --- | --- | --- |
| $AOP_{min}$ | Minimum (diastolic) aortic pressure | 1 | $\pm 10\%$ |
| $AOP_{max}$ | Maximum (systolic) aortic pressure | 1 | $\pm 10\%$ |
| $AOP_{mean}$ | Mean aortic pressure | 1 | $\pm 10\%$ |
| $PAP_{min}$ | Minimum (diastolic) pulmonary arterial pressure | 1 | $\pm 20\%$ |
| $PAP_{max}$ | Maximum (systolic) pulmonary arterial pressure | 1 | $\pm 20\%$ |
| $PAP_{mean}$ | Mean pulmonary arterial pressure | 1 | $\pm 20\%$ |
| $LAP_{mean}$ | Mean left atrial pressure | 1 | $\pm 30\%$ |
| $Qp/Qs$ | pulmonary to systemic flow ratio | 1 | $\pm 20\%$ |
| $PVR_i$ | Indexed PVR | 1 | $\pm 10\%$ |
| $SVR_i$ | Indexed SVR | 1 | $\pm 10\%$ |
| $SV$ | Stroke volume (mL) | 2 | $\pm 20\%$ |
| $EF$ | Ejection fraction (%) | 2 | $\pm 20\%$ |
| $MVPG_{max}$ | Peak mitral valve pressure gradient | 2 | $\pm 20\%$ |
| $AVPG_{max}$ | Peak aortic valve pressure gradient | 2 | $\pm 20\%$ |
| $PVPG_{max}$ | Peak pulmonary valve pressure gradient | 2 | $\pm 20\%$ |
| $VSDPG_{mean}$ | Mean pressure gradient across VSD | 2 | $\pm 20\%$ |
| $ASDPG_{mean}$ | Mean pressure gradient across ASD | 2 | $\pm 20\%$ |
| $PDAPG_{mean}$ | Mean pressure gradient across PDA | 2 | $\pm 20\%$ |

All pressures and pressure gradients are in mmHg. $PVR/SVR$ : WU; $PVR_i/SVR_i$ : $WU \cdot m^2$ .

The areas of the valve annuli (*A_ann_*) are directly incorporated into the model from echo data. For the patients whose echo reports have a qualitative description of valvular stenoses, we assumed stenosis coefficients (*M_st_*) of 0.3 (70% occlusion) and 0.5 (50% occlusion) for severe and moderate stenoses, respectively^35^. The hemodynamic targets used in this study are summarized in **Table 2**, and the raw data for all the patients’ clinical targets are provided in **Supplementary Tables ST2 and ST3**. A comprehensive list of the LPN parameters obtained after tuning is reported in **Supplementary Tables ST4 and ST5** for all ten patients in this study.

### Virtual Surgery

We performed ‘virtual’ surgery on BLV patients by modifying the preoperative LPN model to a BiVR model (**Figure 3A**) and an S1P model (**Figure 3B**). In these figures, we highlighted only the changes compared to the preoperative model (**Figure 1**) as we assumed that the parameters of all other LPN circulation blocks, including the thoracic and the abdominal aorta, upper body, liver, intestines, kidneys, legs, and vena cava, remain the same. To perform a virtual BiVR, we (i) closed the septal defects by removing the resistance between the connected chambers; (ii) repaired and reconstructed the aortic arch by substantially lowering the resistance offered to the blood flow; (iii) performed aortic and/or mitral valve repair by removing the additional resistance, and; (iv) closed the PDA, if present, by removing the resistive element between systemic and pulmonary circulation^47^. Our LPN model for the virtual S1P is identical to that of Corsini et al.^30^. We inserted a 3.5mm BTT shunt between the ascending aorta and pulmonary artery using a nonlinear resistive element calibrated based on the shunt diameter^28^ (**Section B5 in the Supplementary Material**).

**Figure 3.**
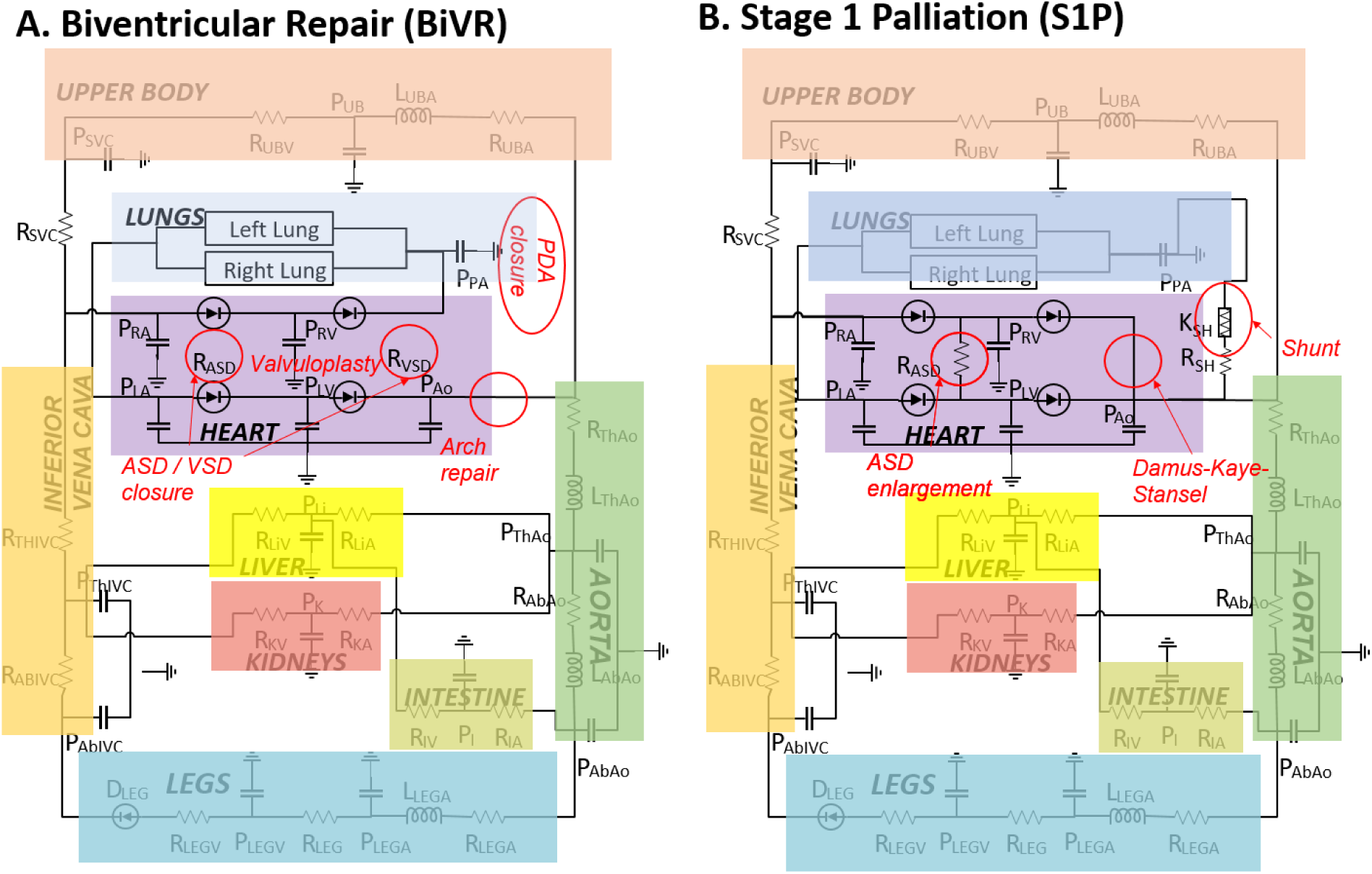
Post-operative LPN models for BLV patients undergoing ‘virtual’ surgery. **(A)** Biventricular repair (BiVR). **(B)** First stage single ventricle palliative procedure (S1P) or Norwood. Virtual repairs performed in each surgical procedure are highlighted. These include **(A)** relief of atrial and ventricular septal defects (ASD, VSD), reconstruction of valve annuli to remove obstruction (valvuloplasty), aortic arch repair, and closure of the patent ductus arteriosus (PDA); **(B)** enlargement of ASD, construction of the Damus-Kaye-Stansel, and inserting a modified 3.5mm Blalock-Taussig-Thomas (BTT) shunt between the systemic and pulmonary circulation systems. *K_SH_*, nonlinear shunt resistance; *R_SH_*, linear resistance of the shunt; All other symbols are described in Figure 1.

### Data Analysis

The Mann-Whitney U test was performed on all continuous variables predicted by the LPN model using MATLAB (Mathworks Inc.). Results are expressed as the mean ± standard deviation. Correlations with a *p*-value < 0.05 were considered statistically significant.

## RESULTS

### Patient Characteristics

The preoperative clinical characteristics of the BLV patients included in this study are provided in **Table 1**. Mean BSA and weight are 0.24±0.03 m^2^ and 4.12±1.70 kg, respectively. The Z-scores of mean MV annulus, LVEDV, and AV annulus are -2.75±1.97, -1.26±2.23, and -3.15±1.77, respectively. All the patients in our cohort had an ASD and a hypoplastic aortic arch, while the majority of the patients had a VSD and mitral and aortic stenoses. Group I (*N*=5) includes patients who underwent the S1P procedure clinically, while Group II (*N*=5) includes patients treated with BiVR. The BiVR procedure consisted of aortic arch repair in 5 patients, ASD closure in 5 patients, and VSD closure in 3 patients. The BSA for these patients ranged between 0.18m^2^ and 0.29m^2^, with no significant differences between Groups I and II. There are no significant differences in the mitral and aortic valves’ Z-scores between the two subgroups (MV Z-score -3.85±2.14 vs. - 1.65±0.88, *p* > 0.1; AV Z-score -3.85±1.57 vs. -2.45±1.67, *p* > 0.1). The measured hemodynamic quantities in which the two groups exhibited significant differences include Q_p_/Q_s_ ratio (1.79±0.73 in Group I vs. 3.44±0.20 in Group II, *p* < 0.01), PAP_mean_ (43.20±9.56 vs. 28.2±7.25), and SVRi (10.76±2.75 vs. 21.66±3.41 WU**·**m^2^, *p* = 0.001).

### Verification of the Preoperative LPN Model

The semi-automatic tuning process applied to the preoperative LPN model resulted in a reasonable agreement between the model predictions and clinical data for the entire patient cohort (**Figure 4**). While **Figure 4** illustrates a graphical comparison of key hemodynamic quantities between the patient’s clinical data and model-predicted values, the raw data are provided in **Supplementary Tables ST6 and ST7** with relative errors quantified. We note that the overall differences are much lower than the standard deviations prescribed during parameter estimation for most clinical measurements, except for a few highlighted quantities in a subset of patients (highlighted in red in **Supplementary Tables ST6 and ST7**). The average percentage error between clinical targets and predicted values is about 9%, well below the prescribed average uncertainty for all targets (∼ 18%).

**Figure 4.**
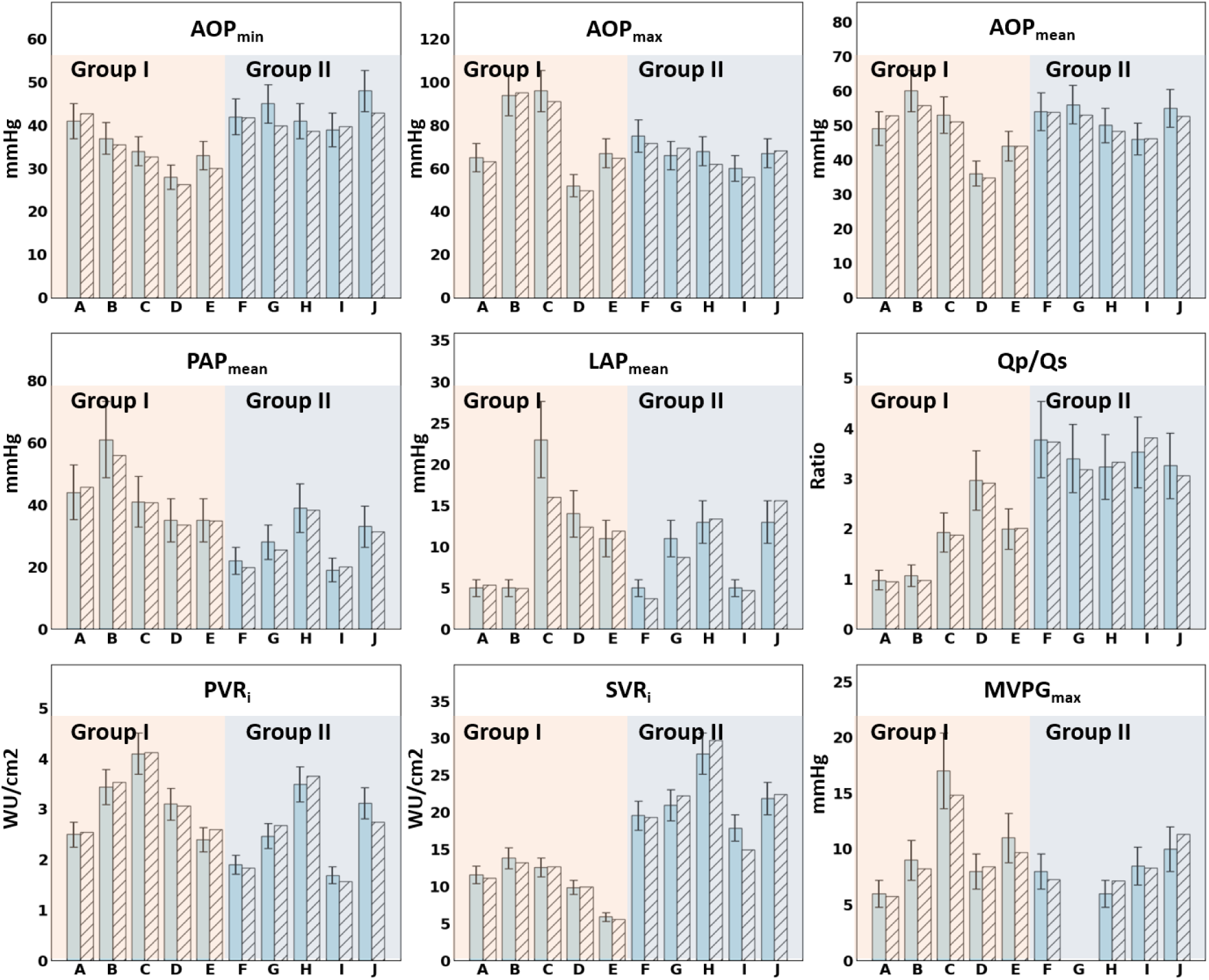
Comparison of optimized LPN-predicted preoperative hemodynamics against clinical data for each patient in the study cohort (A-J), highlighting patient subgroups (Group I, Group II). Group I patients (A-E) were clinically treated with biventricular repair (BiVR), whereas Group II patients (F-J) underwent stage-1 palliation (S1P). Plain bars represent clinical data, while pattern-filled bars represent model predictions. AOP_min/max/mean_: minimum/maximum/mean aortic pressure; PAP_mean_: mean pulmonary artery pressure; LAP_mean_: mean left atrial pressure; Qp/Qs: pulmonary to systemic flow ratio; PVR_i_: indexed pulmonary vascular resistance; SVR_i_: indexed systemic vascular resistance; MVPG_max_: peak mitral valve pressure gradient.

### Virtual Surgery Hemodynamic Outcomes in Group I Patients

The LPN model predicted favorable hemodynamics for a virtual S1P compared to performing a BiVR in Group I patients **(Figure 5**; **Table 3)**. Compared to the preoperative pressures, a virtual S1P on this patient group significantly lowered PAP_max_ (62.57±21.0 vs. 18.50±2.7 mmHg, *p* = 0.01) and PAP_mean_ (42.76±7.7 vs. 17.50±2.7 mmHg, *p* = 0.01). On the contrary, a virtual BiVR didn’t lead to any significant change relative to preoperative values (PAP_max_: 62.57±21.0 vs. 45.40±12.3 mmHg, *p* > 0.1; PAP_mean_: 42.76±7.7 vs. 38.00±10.0 mmHg, *p* > 0.1). PAP_mean_ and PAP_max_ remained significantly higher following a virtual BiVR than a virtual S1P procedure (*p* < 0.05). A substantial increase in LAP_mean_ (10.13±4.3 vs. 25.40±8.2 mmHg, *p* = 0.007) and SVEDP (11.40±4.8 vs. 21.80±8.7 mmHg, *p* = 0.09) is evident after a BiVR, although it did not reach statistical significance for SVEDP. On the other hand, S1P resulted in lowering SVEDP (11.40±4.8 vs. 4.80±1.3 mmHg, *p* < 0.05) and LAP_mean_ (10.13±4.3 vs. 6.20±1.2 mmHg, *p* > 0.1), although the reduction in LAP_mean_ is not statistically significant. However, our model predicted that LAP_mean_ and SVEDP are significantly elevated following a BiVR compared to an S1P in this patient subgroup (*p* < 0.05).

**Figure 5.**
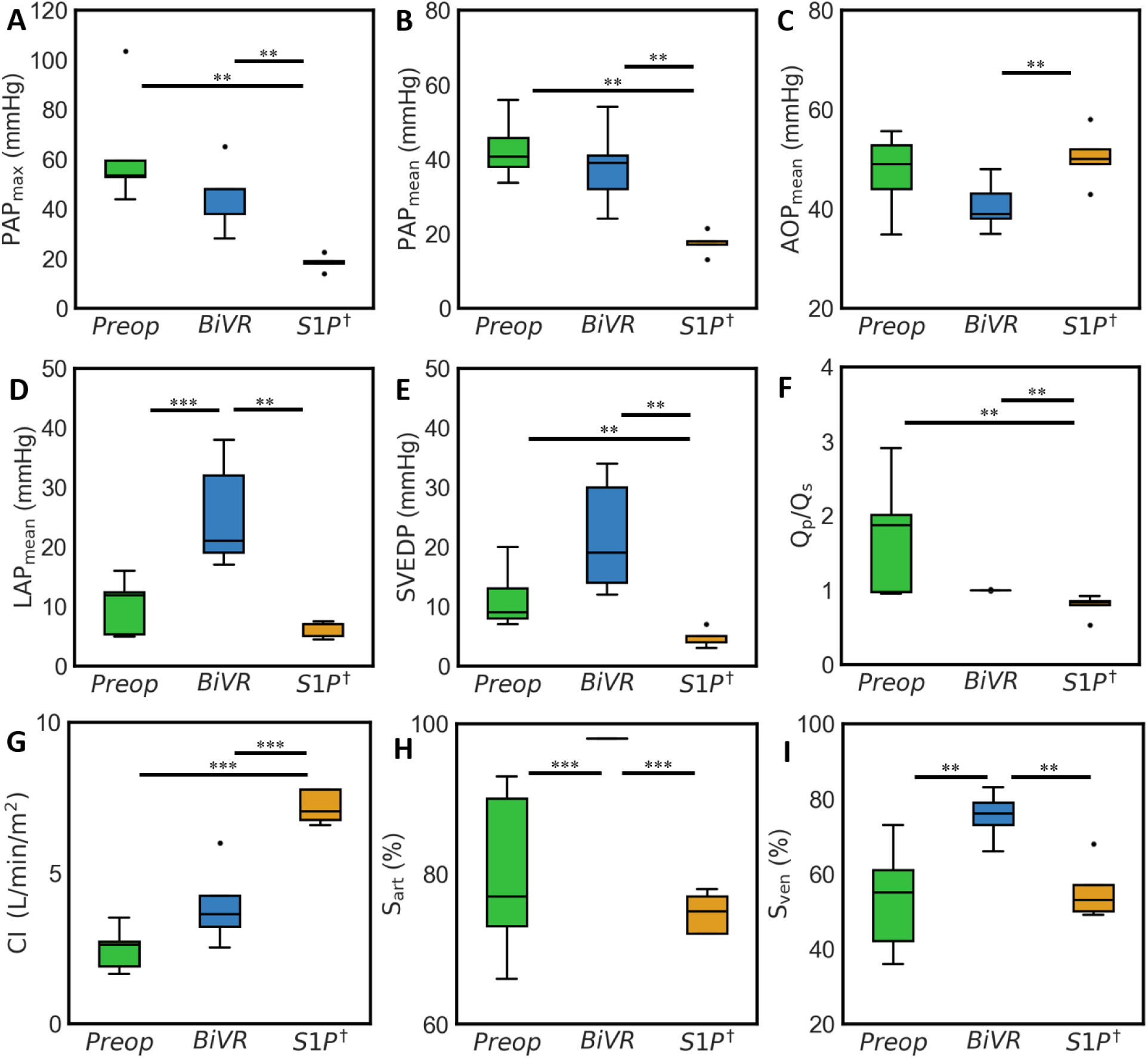
Comparison of predicted preoperative (**preop**) hemodynamic pressures against virtual surgeries (**BiVR**, **S1P**) in Group I (*N*=5) patients. **(A)** max pulmonary arterial pressure (PAP_max_), **(B)** mean pulmonary arterial pressure (PAP_mean_), **(C)** mean aortic pressure (AOP_mean_), **(D)** mean left atrial pressure (LAP_mean_), **(E)** end-diastolic pressure of the systemic ventricle (SVEDP), **(F)** pulmonary-to-systemic flow ratio (Q_p_/Q_s_), **(G)** cardiac index (CI), **(H)** arterial saturation (S_art_), and **(I)** venous saturation (S_ven_). Statistical significance is demonstrated using the Mann-Whitney U test for correlations with *p* < 0.05. Insignificant correlations are not shown. ^†^actual clinical procedure performed on this patient subgroup; \*\**p*<0.05; \*\*\**p*<0.01.

**Table 3.** Predicted hemodynamic variables following virtual BiVR and S1P surgical procedures for Groups I and II patients. The raw data for each individual’s predicted virtual surgery hemodynamics is provided in Supplemental Material.

|  | Group I (N=5) |  |  | Group II (N=5) |  |  |
| --- | --- | --- | --- | --- | --- | --- |
|  | Preop | BiVR | S1P* | Preop | BiVR* | S1P |
| **PAP<sub>max</sub>** | 62.57±21.0 | 45.40±12.3 | 18.50±2.7 | 34.56±8.7 | 20.60±3.1 | 16.50±3.6 |
| **PAP<sub>mean</sub>** | 42.76±7.7 | 38.00±10.0 | 17.50±2.7 | 26.95±7.1 | 15.40±2.8 | 15.40±3.4 |
| **AOP<sub>mean</sub>** | 47.23±7.4 | 40.60±4.5 | 50.40±4.8 | 50.80±3.0 | 74.20±9.1 | 67.40±8.2 |
| **LAP<sub>mean</sub>** | 10.13±4.3 | 25.40±8.2 | 6.20±1.2 | 9.24±4.7 | 6.16±2.5 | 3.20±0.4 |
| **SVEDP** | 11.40±4.8 | 21.80±8.7 | 4.80±1.3 | 9.40±2.6 | 3.60±1.2 | 2.77±0.3 |
| **Qp/Qs** | 1.74±0.7 | 1.00±0.0 | 0.82±0.15 | 3.42±0.3 | 1.00±0.0 | 1.68±0.3 |
| **Qs** | 0.81±0.18 | 0.94±0.19 | 1.12±0.28 | 0.46±0.08 | 0.79±0.16 | 0.69±0.16 |
| **iQs** | 3.31±0.82 | 3.92±1.18 | 4.61±1.54 | 1.89±0.38 | 3.53±0.62 | 3.09±0.59 |
| **CO** | 0.61±0.2 | 0.94±0.2 | 1.94±0.3 | 0.63±0.0 | 0.79±0.2 | 1.79±0.2 |
| **CI** | 2.48±0.65 | 3.92±1.2 | 7.96±1.9 | 2.86±0.35 | 3.53±0.6 | 8.11±1.0 |
| **S<sub>art</sub> (%)** | 79.80±10.2 | 98.00±0.0 | 74.80±2.5 | 92.4±4.5 | 98.00±0.0 | 82.20±1.7 |
| **S<sub>ven</sub> (%)** | 53.40±13.25 | 75.40±5.8 | 55.40±6.9 | 45.80±9.74 | 74.80±5.5 | 55.00±7.6 |
| **O<sub>2</sub>D** | 541.00±112.3 | 816.80±240.4 | 722.80±277.8 | 412.00±90.0 | 743.20±128.7 | 544.40±99.1 |

\*indicates the clinically performed procedure for the particular group. All pressures are in mmHg. Qs: Liters/min; iQs: L/min/m<sup>2</sup>; CO: Liters/min; CI: L/min/m<sup>2</sup>; O<sub>2</sub>D: mlO<sub>2</sub>/min/m<sup>2</sup>

The flow measures and oxygen saturations predicted by the model are consistent with the physiology of BiVR and S1P procedures **(Figure 5**; **Table 3)**. Notably, CI is significantly lowered following a virtual BiVR in these Group I patients than after a virtual S1P (7.96±1.9 vs. 3.92±1.2 L/min/m^2^, *p* < 0.01). While BiVR resulted in evenly distributed Q_p_/Q_s_ (1.00±0.0), the ratio is significantly reduced after S1P compared to the preoperative flow ratio (1.74±0.7 vs. 0.82±0.15, *p* < 0.05). Likewise, oxygen saturations S_art_ and S_ven_ are significantly higher after a BiVR compared to S1P (S_art_: 98.00±0.0 vs. 74.80±2.5 %, *p* = 0.007; S_ven_: 75.4±5.80 vs. 55.40±6.90 %, *p* = 0.015). Overall, the model-predicted hemodynamics favored S1P over BiVR for Group I patients, retrospectively corroborating the clinically performed procedure as a BiVR in this group of patients would lead to a non-sustainable physiology with hypertension in LA and PA. The raw data for each patient’s predicted virtual surgery hemodynamics in this subgroup are provided in **Supplementary Table ST8**.

### Virtual Surgery Hemodynamic Outcomes in Group II Patients

The LPN model didn’t predict any adverse hemodynamic outcome after a virtual BiVR in Group II patients, retrospectively corroborating the clinically performed BiVR procedure in this patient subgroup **(Figure 6**; **Table 3)**. Compared to preoperative values, both surgical approaches showed a significant decrease in PAP_max_ (BiVR: 34.56±8.7 vs. 20.60±3.1 mmHg, *p* = 0.02; S1P: 34.56±8.7 vs. 16.50±3.6 mmHg, *p* = 0.01) and PAP_mean_ (BiVR: 26.95±7.1 vs. 15.40±2.8 mmHg, *p* = 0.02; S1P: 26.95±7.1 vs. 15.40±3.4 mmHg, *p* = 0.01) and a significant increase in AOP_mean_ (BiVR: 50.80±3.0 vs. 74.20±9.1 mmHg, *p* < 0.01; S1P: 50.80±3.0 vs. 67.40±8.2 mmHg, *p* < 0.05). Contrary to Group I, performing a virtual BiVR in Group II did not lead to LA hypertension or high SVEDP (LAP_mean_: 9.24±4.7 vs. 6.16±2.5 mmHg, *p* > 0.1; SVEDP: 9.40±2.6 vs. 3.60±1.0 mmHg preoperatively, *p* = 0.01). PAP, LAP, and SVEDP are significantly reduced after a virtual BiVR in Group I compared to Group II (PAP_max_: 45.40±12.3 vs. 20.60±3.1 mmHg, *p*=0.01; PAP_mean_: 38.00±10.0 vs. 15.40±2.8 mmHg, *p*=0.01; LAP_mean_: 25.40±8.2 vs. 6.16±2.5 mmHg, *p*=0.007; SVEDP: 21.80±8.7 vs. 3.60±1.2 mmHg, *p*=0.012).

**Figure 6.**
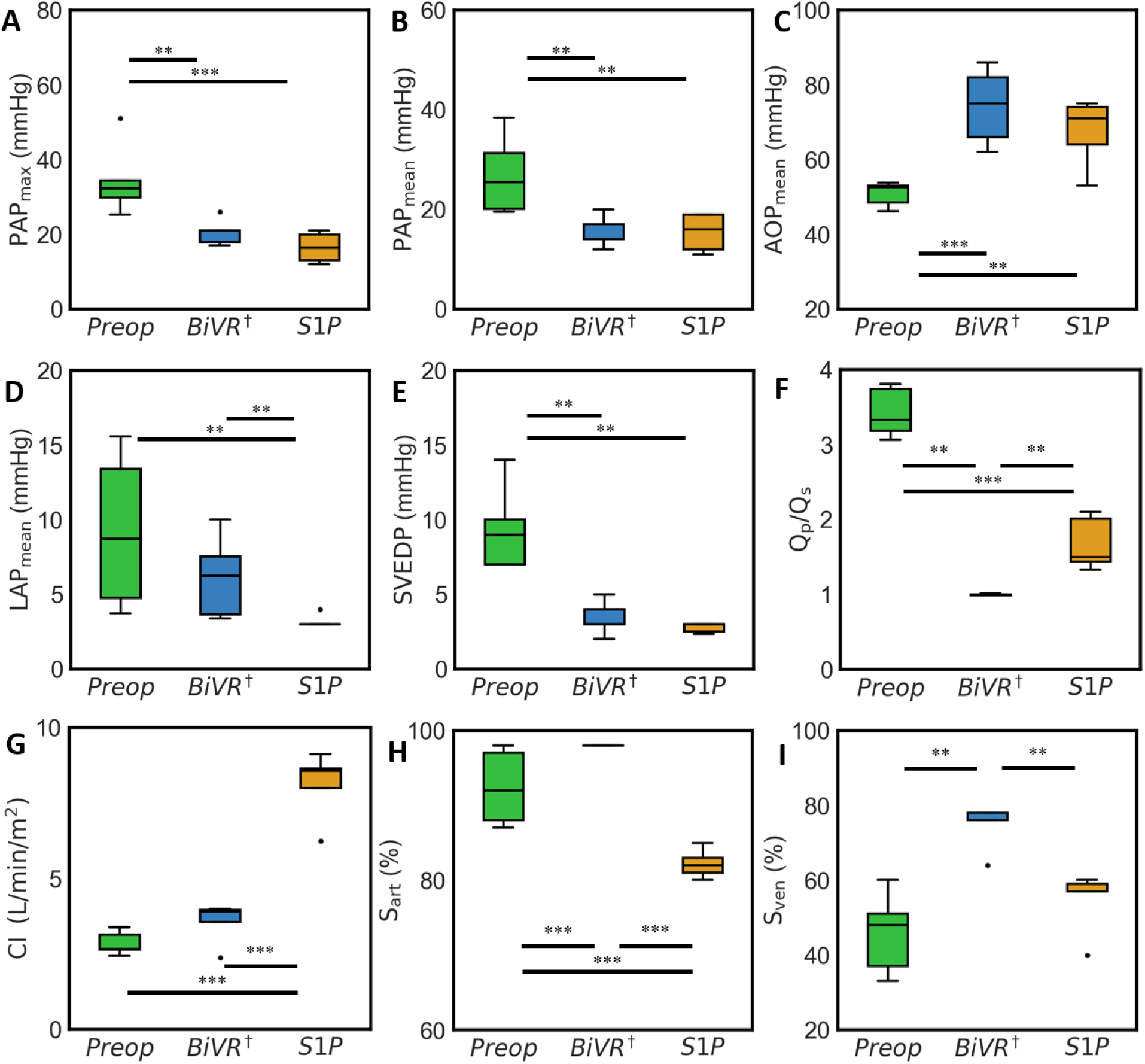
Comparison of predicted preoperative (**preop**) hemodynamic pressures against virtual surgeries (**BiVR**, **S1P**) in Group II (*N*=5) patients. **(A)** max pulmonary arterial pressure (PAP_max_), **(B)** mean pulmonary arterial pressure (PAP_mean_), **(C)** mean aortic pressure (AOP_mean_), **(D)** mean left atrial pressure (LAP_mean_), **(E)** end-diastolic pressure of the systemic ventricle (SVEDP), **(F)** pulmonary-to-systemic flow ratio (Q_p_/Q_s_), **(G)** cardiac index (CI), **(H)** arterial saturation (S_art_), and **(I)** venous saturation (S_ven_). Statistical significance is demonstrated using the Mann-Whitney U test for correlations with *p* < 0.05. Insignificant correlations are not shown. ^†^actual clinical procedure performed on this patient subgroup; \*\**p*<0.05; \*\*\**p*<0.01.

Similar to Group I patients, the flow rates and oxygen saturations predicted in Group II are consistent with the physiologies of BiVR and S1P and conform to the patient characteristics of this group (**Figure 6**, **Table 3**). The balance in Q_p_/Q_s_ is restored after a BiVR (1.00±0.0), while a virtual S1P had a higher Q_p_/Q_s_ ratio (1.68±0.3 vs. 1.00+0.0, *p* < 0.05). Consistent with S1P physiology, where the systemic ventricle supports pulmonary circulation, CI is significantly higher after a virtual S1P than a virtual BiVR (3.53±0.6 vs. 8.11±1.0, *p* < 0.01). Likewise, oxygen saturations S_art_ and S_ven_ are significantly higher after a BiVR compared to S1P (S_art_: 98.00±0.0 vs. 82.20±1.7 %, *p* = 0.007; S_ven_: 74.80±5.5 vs. 55.00±7.6 %, *p* = 0.01). Overall, we demonstrated that performing virtual BiVR on Group II patients will not lead to high LA or PA pressures, thus supporting retrospectively the validity of the clinically performed procedure in this patient subgroup. The raw data for each patient’s predicted virtual surgery hemodynamics in this subgroup are provided in **Supplementary Table ST9**.

### Comparison Against Post-Operative Cath Data

We compared the LPN model-predicted hemodynamic pressures (AOP_mean_, PAP_mean_) and flow indices (Qp, Qs, Qp/Qs, and CI) against post-operative cath measurements in a subset of Group I patients **(Table 4)**. Patients who did not have post-operative data were not included in this comparison. Group II patients were also excluded as post-operative cath-based evaluation is not part of their clinical routine. These measurements were clinically obtained before performing Stage 2 palliation, i.e., the Glenn procedure. While the relative errors are reasonably below 10% in Patient A for all the quantities, the relative errors are marginally higher than the 10% threshold in Patient D. Specifically, in Patient A, the model predicted AOP_mean_, PAP_mean_, and Qs exhibited a strong agreement with cath data with relative errors below 3%, while CI has a slightly elevated difference (5.8%) and Qp and Qp/Qs ratio differed from cath data by a reasonable 10%. However, in Patient D, the relative errors between the LPN model and cath data in AOP_mean_ and Qs are within 5%, while CI has a marginally higher relative error (5.4%). However, the relative errors in PAP_mean_, Qp, and Qp/Qs ratio are slightly above 10%, as highlighted in **Table 4**.

**Table 4.** Comparison of model-predicted post-operative hemodynamic quantities against catheterization measurements in a subset of Group I patients. The error in LPN prediction, measured as % of the clinical data, is shown in parentheses.

| Patient | A |  |  | D |  |  |
| --- | --- | --- | --- | --- | --- | --- |
|  | Cath | Model (as-is) | Model (BSA-scaled) | Cath | Model (as-is) | Model (BSA-scaled) |
| **AOP<sub>mean</sub> (mmHg)** | 49.0 | 49.0 (0.0%) | 49.8 (1.6%) | 41.0 | 43.0 (4.9%) | 41.8 (2.0%) |
| **PAP<sub>mean</sub> (mmHg)** | 13.3 | 13.0 (2.3%) | 12.6 (5.3%) | 16.0 | 18.0 (12.5%) | 14.4 (10.0%) |
| **Qp (L/min)** | 0.80 | 0.88 (10.0%) | 0.83 (3.8%) | 0.54 | 0.60 (11.0%) | 0.54 (0.0%) |
| **Qs (L/min)** | 1.05 | 1.07 (2.0%) | 1.10 (4.8%) | 0.74 | 0.75 (2.0%) | 0.76 (2.7%) |
| **Qp/Qs** | 0.76 | 0.84 (10.0%) | 0.75 (1.3%) | 0.72 | 0.80 (11.0%) | 0.71 (1.4%) |
| **CI (L/min)** | 6.38 | 6.75 (5.8%) | 6.22 (2.5%) | 6.10 | 6.43 (5.4%) | 6.19 (1.5%) |

\*Both patients underwent S1P and were examined before Stage 2 palliation (i.e., Glenn procedure).

To account for the patient’s growth from Stage 1 to Stage 2 palliation, we have performed additional simulations where the S1P model parameters are adjusted by the change in the patient’s BSA. We notice a reasonable improvement in the comparison against cath data, where all the model-predicted hemodynamic quantities are within a 5% margin compared to cath measurements (column BSA-scaled in **Table 4**). Only patient D’s PAP_mean_ differed by 10% compared to cath data (16 mmHg from cath vs. 14.4 mmHg predicted by the BSA-scaled model).

## DISCUSSION

In this study, we innovated a patient-specific data-driven modeling framework to perform virtual surgery in BLV neonates with underdeveloped left-sided heart structures. We demonstrated that the *digital twin* predictions endorse the clinically performed surgery in a small, retrospectively identified patient cohort (*N*=10). In particular, in the subgroup of patients who clinically underwent S1P (Group I, *N*=5), the model predicted adverse hemodynamics after a virtual BiVR with significantly high left atrial and pulmonary artery pressures. Likewise, in the patient subgroup who were offered a BiVR (Group II, *N*=5), the model didn’t anticipate any alarming hemodynamic outcomes that would favor an S1P instead. These results suggest that the current patient-specific predictive modeling framework may potentially guide clinicians in making personalized decisions between BiVR and S1P in BLV patients.

### Motivation to employ predictive computational models

BLV patients form a spectrum between a nearly normal cardiac structure and a congenital disability leading to a severely underdeveloped hypoplastic left heart, HLHS. While there is no ambiguity in treating HLHS patients who typically undergo the multi-staged single ventricle palliative procedure, pediatric surgeons often face a critical dilemma in treating BLV patients due to the usually dichotomous decision-making during the neonatal period, whether to perform a BiVR or an S1P^2^. Pediatric surgeons may often consider delaying this climacteric two-pronged decision-making to a later stage by performing an interim palliative procedure, restoring blood flow through the left heart and allowing the structures to grow, thereby mitigating operative challenges and allowing the left heart to be gradually exposed to systemic loads^10–14^. This staging is typically adopted in patients born with endocardial fibroelastosis (EFE) or deemed at high risk for cardiopulmonary bypass as newborns. However, despite promising on its surface, the approach found limited clinical success (∼33%)^15,16^. Moreover, the lack of predictive capabilities in the current clinical workflow may eventually lead to a discordant pursuit of BiVR, increasing the risk of operative mortality.

Current size-based metrics for diagnosis and treatment planning make it challenging to predict the outcome of a BiVR until the surgery is attempted^23^. Further, switching a failed two-ventricle surgery to a univentricular circuit is fraught with risk and high mortality^2^. On the contrary, if a BLV patient undergoes the S1P procedure, there is no way to confirm the likelihood of a successful BiVR. Our BLV digital twin may address these critical issues in pediatric heart surgery by enabling pediatric cardiologists and thoracic surgeons to predict post-operative hemodynamics, including pressures and flow distributions, make a quantitative comparison between the two surgical pathways, and choose the hemodynamically optimal procedure, before operating on the patient.

### Robustness of the LPN-based model and semi-automatic method for parameter estimation

Our LPN-based computational model for BLV patients was adapted from a well-established model for S1P circulation^30,48^. LPN-based models are known to be superior at rapidly and effectively understanding the organ-level and system-level hemodynamic impact^33,48^, including capturing ventricular end-diastolic and end-systolic pressure-volume relations (EDPVR, ESPVR)^49,50^, sarcomere kinematics and adaptation^51^, remodeling due to systolic and diastolic dysfunction and valvular stenoses and regurgitation^49^, unlike the expensive computational fluid dynamics models^33,52,53^. However, a challenge in their large-scale adoption is the relatively large number of parameters that must be tuned for personalization, posing identifiability issues^54^. Recent advances in parameter estimation methods employing Bayesian statistics have improved the robustness of the tuning process and reduced the subjectivity^38^. We have leveraged these advances to develop a semi-automatic tuning framework that personalizes the preoperative LPN model for BLV patients. The optimization process for tuning the LPN model takes about three hours for each BLV patient while obtaining a reasonable agreement with the clinical data (**Figure 4, Tables ST6 and ST7 in the Supplementary Material**) and could be quickly adopted into the clinical workflow^48^ A few exceptions with substantial discrepancies are the transvalvular pressure gradients and other hemodynamic quantities derived from echo measurements, known to be noise-sensitive. Moreover, these higher discrepancies are not unexpected as they are associated with a relatively higher standard deviation during parameter estimation (see Step 3 of the LPN Parameter Estimation subsection in Methods).

Using the personalized preoperative LPN model as the baseline, we performed virtual surgery simulations of BiVR and S1P on every BLV patient in the cohort and compared the model-predicted hemodynamics to identify the most favorable procedure. In performing the virtual surgery, our zero-dimensional (0D) LPN model only accounts for the functional impact of the surgical operation and not the actual change in the morphology or the structure. For instance, when reconstructing the hypoplastic aortic arch, we lower the resistance to the blood flow iteratively until the pressure gradient across the constricted arch reduces to at most a few millimeters of Hg. Likewise, during valvuloplasty, we reset the stenosis parameter (*M_st_*) in our versatile valve model to the unstenosed state that virtually removes any excess blockage to the flow through the valvular orifice^35^. Although prior studies have explored using LPN models to study ventricular remodeling due to stenoses and regurgitation^49,55^, we did not explicitly analyze these effects in this study. The robustness of the LPN model and its utility for predictive modeling is also demonstrated by obtaining a reasonable agreement against available post-operative cath data in a subset of Group I patients. We note that this post-operative clinical data was obtained a few months later, before the Glenn procedure, and likely contributed to the modest discrepancies observed in some quantities when the comparison is made ‘as-is’ (**Table T4**). However, when the model parameters are scaled by the change in the patient’s BSA, accounting for the patient’s growth, we noticed a more reasonable agreement between model predictions and cath data.

### Hemodynamic assessment of virtual surgery predictions

Our central premise for this study is that patient-specific predictive computational modeling could be employed to determine the hemodynamically most favorable surgical path in BLV patients, and it was tested in a small, retrospectively identified patient cohort. In Group I, who clinically underwent an S1P, performing a BiVR would have been detrimental as it would have led to a significant increase in LAP and SVEDP compared to the preoperative values while only marginally affecting PAP, which is already high (**Figure 5, Supplementary Table ST8**). At the same time, the model predicts that an S1P in this patient group would be more favorable as it significantly lowers PAP, LAP, and SVEDP to nominal values for these patients. This prediction corroborates retrospectively the choice made by the multi-disciplinary clinical team, although without the ability to incorporate the forecasted hemodynamic outcome into their decision-making. While the model predicted flow distributions (Q_p_/Q_s_, CI) and oxygen saturations (S_art_, S_ven_) and delivery (O_2_D) are consistent with the respective physiology and are reasonably within the physiological values for this group for both the virtual surgeries (**Figure 5**), it is not immediately apparent how we could incorporate this data into clinical decision-making, warranting further research.

For Group II patients, the model predictions recommend that BiVR is the most appropriate choice, corroborating retrospectively the choice made by the multi-disciplinary clinical team. In this case, however, if we compare the model-predicted pressures between the two virtual surgeries, we may find S1P to be marginally better than BiVR, particularly in lowering the PAP, LAP, and SVEDP relative to preoperative values (**Figure 6, Supplementary Table ST9**). Nonetheless, we also find that the absolute values of these pressures following a BiVR are not adverse to choosing S1P. Moreover, an S1P on these patients will lead to poor O_2_ saturation and delivery compared to performing a BiVR (**Figure 6, Supplementary Table ST9**) and may increase the risk of encountering liver disease^5^. Therefore, when applying our digital twin for clinical decision-making, one should assess whether the LAP, PAP, and SVEDP after a virtual BiVR are within acceptable limits to justify the benefits of a two-ventricle physiology and avoid the deleterious long-term consequences of the Fontan physiology^4^.

Although we have analyzed only a few hemodynamic quantities of clinical interest, such as cardiac output and mean and min/max pressures, the LPN model can provide detailed ventricular pressure-volume loops, used to evaluate changes in cardiac work and mechanical efficiency, and time-dependent pressure and flow waveforms (**Figures SF1-SF3 in the Supplementary Material**). In this pilot study, cardiac stroke work (area under the ventricular pressure-volume diagram, **Supplemental Figure SF1**) is usually higher after an S1P than BiVR for all the patients. We attribute this to the increased load on the systemic right ventricle supporting both pulmonary (through BTT shunt) and systemic circulations. Therefore, although stroke work and its derivatives have been shown to be key biomarkers in other studies^34,56–59^, further research is required to understand their effectiveness in choosing between a BiVR and S1P for BLV patients.

### Limitations

We recognize several limitations of our study. Our 0D LPN model doesn’t account for the complex geometrical structure and morphology of the malformed atria and ventricles in these BLV patients. Instead, only the integral measures, such as the cavity volumes and functional hemodynamic impact modeled using complex resistive elements, are included. Moreover, due to its low fidelity, the model cannot resolve detailed spatial dynamics, such as blood velocity maps, flow pressure distributions, tissue stresses, or strain profiles, which require complex multiscale fluid and solid mechanics models^33,52,53,60–66^. In applications where such a detailed analysis is not required due to a lack of adequate imaging data or limited time for decision-making to perform complex computations, LPN models are helpful tools that will provide key hemodynamic data in a clinically relevant timeframe and may assist decision-making^48^. Moreover, LPN model computations are fast, do not require extensive computing infrastructure, and are versatile to propagate uncertainty in the input data to estimate confidence intervals on the predicted post-operative hemodynamics^36,48,67^. Our LPN model also closely resembles the one developed by Broome et al.^49^, employing a time-varying elastance function for ventricular pressure-volume (PV) relation and a versatile valve model, known to produce realistic PV loops for varying preloads^49^. However, although the PV relation is assumed to be the same across all the patients, the parameters in this relation, including myocardial resting volume (V_0_), passive stiffness (*c*, *d*), and active elastance parameters (*a*, *b*), are tuned to match the patient’s stroke volume (SV), ejection fraction (EF), and aortic pressures (AOP_min/max/mean_), along with other available clinical data (**Table 2**). Therefore, our model produces unique PV loops for each patient (**Figure SF1 in Supplementary Material**). Nevertheless, we did not analyze the patient’s preload-recruitable stroke work (PRSW)^50^ or any other elastance metrics based on the EDPVR or ESPVR, as this data is not available clinically for each patient and is beyond the scope of this work^49,50^.

In the current study, we assume that the LPN parameters of the post-virtual surgery models remain the same as those of the preoperative model. This is a reasonable assumption as we are interested in predicting only the immediate post-operative outcomes to support clinical decision-making by providing additional hemodynamic data, which is lacking. However, our model does not account for myocardial remodeling due to the relief of valvular obstruction, transient changes in ventricular performance and loading during the surgery, long-term remodeling due to pressure and volume overloading, and other auto-regulatory mechanisms. Although we demonstrated statistical significance in our data, the patient cohort in this pilot study is small (*N*=10) and is only retrospective. Moreover, the current cohort does not include challenging patients with controversial decision-making or failed initial surgery, prompting them to be switched to an alternative pathway.

In a follow-up study, we will validate the model on a more extensive and prospective patient group, including validating the predicted immediate post-operative hemodynamics using cath and echo data. We will further challenge the model to evaluate its performance on controversial patients with failed BiVR. We will also account for uncertainties in the clinical measurements and evaluate the sensitivities of the LPN parameters and their effect on predicted hemodynamics^38,67^. We will also explore the model’s applicability to other palliative surgical pathways for BLV patients, including the hybrid approach or the staged LV recruitment procedure^10,13^, and account for ventricular growth between the procedures^68^.

## CONCLUSIONS

We developed a novel lumped parameter network (LPN)-based model of the circulatory system for patients born with a borderline left ventricle (BLV). The LPN model was personalized to a patient’s cath and echo data using a semi-automatic tuning framework and was modified to simulate biventricular repair (BiVR) and stage-1 palliation (S1P or Norwood) surgical procedures. The model was applied retrospectively to a small BLV patient cohort (*N*=10). Virtual surgeries (BiVR and S1P) were performed on each patient to predict hemodynamics after each surgery, and they were compared to determine the hemodynamically most favorable pathway. We demonstrated that the model predictions from this bespoke digital twinning framework endorsed the clinical decision made in both BiVR and S1P groups and may potentially guide clinicians in making a simulation-informed decision between BiVR and S1P in this critical patient group. While the current modeling framework holds promise, it must be validated on a larger and prospective patient cohort, including challenging and controversial cases with a failed initial pathway.

## GLOSSARY OF ABBREVIATIONS

AOP: Aortic pressure
ASD: Atrial septal defect
ASDPG: Pressure gradient across the atrial septal defect
AV: Aortic valve
AVPG: Aortic valve pressure gradient
BiVR: Biventricular repair
BLV: Borderline left ventricle
BSA: Body Surface Area
BTTs: Blalock-Taussig Thomas shunt
CO: Cardiac output
CV_O2_: Whole-body O_2_ consumption
CVP: Central venous pressure
EDP: End-diastolic pressure
EDV: End-diastolic volume
EF: Ejection Fraction
HLHS: Hypoplastic left heart syndrome
LA: Left atrium
LAP: Left atrial pressure
LAH: Left atrial hypertension
LPN: Lumped parameter network
LV: Left ventricle
LVOT: Left ventricular outflow tract
MV: Mitral valve
MVPG: Pressure gradient across the mitral valve
O_2_D: Oxygen delivery
PA: Pulmonary artery
PAB: Pulmonary artery banding
PAP: Pulmonary arterial pressure
PDA: Patent ductus arteriosus
PDAPG: Pressure gradient across the patent ductus arteriosus
PVPG: Pulmonary valve pressure gradient
PVR: Pulmonary vascular resistance
PVRi: Indexed PVR
Qp: Pulmonary flow rate
Qs: Systemic flow rate
RA: Right atrium
RAP: Right atrial pressure
RCR: Resistor-capacitor-resistor
RV: Right ventricle
RVOT: Right ventricular outflow tract
S_art_: Arterial saturation
SV: Stroke volume
SVCP: Superior vena cava pressure
SVEDP: Systemic ventricular end-diastolic pressure
SVEDV: Systemic ventricular end-diastolic volume
S_ven_: Venous saturation
S1P: Stage 1 palliation
SVR: Systemic vascular resistance
SVRi: Indexed SVR
VSD: Ventricular septal defect
VSDPG: Pressure gradient across the ventricular septal defect
VTI: Velocity time integral

## DECLARATIONS

### Ethics Statement

This single-center study was performed following HIPAA-compliant processes and guidelines and was approved by the Columbia University Irving Medical Center Institutional Review Board (IRB#AAAT2664 approved on July 10, 2020), while obtaining informed consent was waived due to the study being retrospective.

### Consent for Publication

All authors have agreed with the content and given explicit consent to submit the manuscript for publication.

### Data Availability

Data will be shared upon a reasonable request from the corresponding authors.

### Authors’ Contributions

**Yurui Chen** (Data curation; Formal analysis; Investigation; Methodology;

Software; Validation; Visualization; Writing – original draft; Writing – review & editing)

**Isao A. Anzai** (Data curation; Formal analysis; Writing – review & editing)

**David M. Kalfa** (Conceptualization; Data curation; Formal analysis; Funding acquisition; Investigation; Project administration; Supervision; Validation; Writing – review & editing)

**Vijay Vedula** (Conceptualization; Data curation; Formal analysis; Funding acquisition; Investigation; Methodology; Project administration; Resources; Software; Supervision; Validation; Visualization; Writing – original draft; Writing – review & editing)

## Acknowledgments

YC and VV would like to acknowledge helpful discussions with Dr. Justin Tran on the parameter estimation framework for lumped parameter networks.

## REFERENCES

1. Corno AF. Borderline left ventricle. European Journal of Cardio-thoracic Surgery. 2005;27(1):67–73. doi:10.1016/j.ejcts.2004.10.034

2. Cohen MS. Assessing the borderline ventricle in a term infant: Combining imaging and physiology to establish the right course. Curr Opin Cardiol. 2018;33(1):95–100. doi:10.1097/HCO.0000000000000466

3. De Leval MR. The Fontan circulation: What have we learned? What to expect? Pediatr Cardiol. 1998;19(4):316–320. doi:10.1007/s002469900315

4. Rychik J. The Relentless Effects of the Fontan Paradox. Semin Thorac Cardiovasc Surg Pediatr Card Surg Annu. 2016;19(1):37–43. doi:10.1053/j.pcsu.2015.11.006

5. Ghbeis MB, Pane C, Beroukhim R, et al. Biventricular Repair of Univentricular Heart Lowers Risk of Liver Disease Compared With the Fontan Operation. JACC: Advances. 2025;4(1):1–10. doi:10.1016/j.jacadv.2024.101429

6. Feinstein JA, Benson DW, Dubin AM, et al. Hypoplastic left heart syndrome: Current considerations and expectations. J Am Coll Cardiol. 2012;59(1 SUPPL.):S1-S42. doi:10.1016/j.jacc.2011.09.022

7. Ghanayem NS, Allen KR, Tabbutt S, et al. Interstage mortality after the Norwood procedure: Results of the multicenter Single Ventricle Reconstruction trial. Journal of Thoracic and Cardiovascular Surgery. 2012;144(4):896–906. doi:10.1016/j.jtcvs.2012.05.020

8. Karamlou T, Overman D, Hill KD, et al. Stage 1 hybrid palliation for hypoplastic left heart syndrome-assessment of contemporary patterns of use: An analysis of The Society of Thoracic Surgeons Congenital Heart Surgery Database. Journal of Thoracic and Cardiovascular Surgery. 2015;149(1):195–202.e1. doi:10.1016/j.jtcvs.2014.08.020

9. Schwartz ML, Gauvreau K, Geva T. Predictors of Outcome of Biventricular Repair in Infants With Multiple Left Heart Obstructive Lesions. Circulation. 2001;104:682–687. doi:10.1007/s00246-011-0142-2

10. Haller C, Honjo O, Caldarone CA, Van Arsdell GS. Growing the Borderline Hypoplastic Left Ventricle: Hybrid Approach. Operative Techniques in Thoracic and Cardiovascular Surgery. 2016;21(2):124–138. doi:10.1053/j.optechstcvs.2017.03.001

11. Ballard G, Tibby S, Miller O, et al. Growth of left heart structures following the hybrid procedure for borderline hypoplastic left heart. European Journal of Echocardiography. 2010;11(10):870–874. doi:10.1093/ejechocard/jeq085

12. Emani SM, Bacha EA, McElhinney DB, et al. Primary left ventricular rehabilitation is effective in maintaining two-ventricle physiology in the borderline left heart. Journal of Thoracic and Cardiovascular Surgery. 2009;138(6):1276–1282. doi:10.1016/j.jtcvs.2009.08.009

13. Emani SM, McElhinney DB, Tworetzky W, et al. Staged left ventricular recruitment after single-ventricle palliation in patients with borderline left heart hypoplasia. J Am Coll Cardiol. 2012;60(19):1966–1974. doi:10.1016/j.jacc.2012.07.041

14. Emani SM. Staged Left Ventricular Recruitment and Biventricular Conversion for Patients With Borderline Left Heart. Operative Techniques in Thoracic and Cardiovascular Surgery. 2016;21(2):112–123. doi:10.1053/j.optechstcvs.2017.02.003

15. Herrin MA, Zurakowski D, Baird CW, et al. Hemodynamic parameters predict adverse outcomes following biventricular conversion with single-ventricle palliation takedown. Journal of Thoracic and Cardiovascular Surgery. 2017;154(2):572–582. doi:10.1016/j.jtcvs.2017.02.070

16. Chiu P, Emani S. Left Ventricular Recruitment in Patients With Hypoplastic Left Heart Syndrome. Semin Thorac Cardiovasc Surg Pediatr Card Surg Annu. 2021;24:30–36. doi:10.1053/j.pcsu.2021.03.001

17. Plymale JM, Frommelt PC, Nugent M, Simpson P, Tweddell JS, Shillingford AJ. The Infant with Aortic Arch Hypoplasia and Small Left Heart Structures: Echocardiographic Indices of Mitral and Aortic Hypoplasia Predicting Successful Biventricular Repair. Pediatr Cardiol. 2017;38(6):1296–1304. doi:10.1007/s00246-017-1661-2

18. Szwast AL, Marino BS, Rychik J, Gaynor JW, Spray TL, Cohen MS. Usefulness of left ventricular inflow index to predict successful biventricular repair in right-dominant unbalanced atrioventricular canal. American Journal of Cardiology. 2011;107(1):103–109. doi:10.1016/j.amjcard.2010.08.052

19. Mart CR, Eckhauser AW. Development of an Echocardiographic Scoring System to Predict Biventricular Repair in Neonatal Hypoplastic Left Heart Complex. Pediatr Cardiol. 2014;35(8):1456–1466. doi:10.1007/s00246-014-1009-0

20. Tuo G, Khambadkone S, Tann O, et al. Obstructive left heart disease in neonates with a “borderline” left ventricle: Diagnostic challenges to choosing the best outcome. Pediatr Cardiol. 2013;34(7):1567–1576. doi:10.1007/s00246-013-0685-5

21. Kovalchin JP, Brook MM, Rosenthal GL, Suda K, Hoffman JI e., Silverman NH. Echocardiographic hemodynamic and morphometric predictors of survival after two-ventricle repair in infants with critical aortic stenosis. J Am Coll Cardiol. 1998;32(1):237–244. doi:10.1016/S0735-1097(98)00218-6

22. Parsons MK, Moreau GA, Graham TP, Johns JA, Boucek RJ. Echocardiographic estimation of critical left ventricular size in infants with isolated aortic valve stenosis. J Am Coll Cardiol. 1991;18(4):1049–1055. doi:10.1016/0735-1097(91)90765-2

23. Cantinotti M, Marchese P, Giordano R, et al. Echocardiographic scores for biventricular repair risk prediction of congenital heart disease with borderline left ventricle: a review. Heart Fail Rev. 2022;(0123456789). doi:10.1007/s10741-022-10230-0

24. Mart CR, Eckhauser AW. Development of an Echocardiographic Scoring System to Predict Biventricular Repair in Neonatal Hypoplastic Left Heart Complex. Pediatr Cardiol. 2014;35(8):1456–1466. doi:10.1007/s00246-014-1009-0

25. Lofland GK, McCrindle BW, Williams WG, et al. Critical aortic stenosis in the neonate: A multi-institutional study of management, outcomes, and risk factors. Journal of Thoracic and Cardiovascular Surgery. 2001;121(1):10–27. doi:10.1067/mtc.2001.111207

26. Cohen MS, Jacobs ML, Weinberg PM, Rychik J. Morphometric analysis of unbalanced common atrioventricular canal using two-dimensional echocardiography. J Am Coll Cardiol. 1996;28(4):1017–1023. doi:10.1016/S0735-1097(96)00262-8

27. Rhodes LA, Colan StevenD, Perry SB, Jonas RA, Sanders SP. Predictors of survival in neonates with critical aortic stenosis. Circulation. 1991;84:2325–2335.

28. Migliavacca F, Pennati G, Dubini G, et al. Modeling of the Norwood circulation: Effects of shunt size, vascular resistances, and heart rate. Am J Physiol Heart Circ Physiol. 2001;280(5 49-5):2076-2086. doi:10.1152/ajpheart.2001.280.5.h2076

29. Migliavacca F, Dubini G, Pennati G, et al. Computational model of the fluid dynamics in systemic-to-pulmonary shunts. J Biomech. 2000;33(5):549–557. doi:10.1016/S0021-9290(99)00219-5

30. Corsini C, Baker C, Kung E, et al. An integrated approach to patient-specific predictive modeling for single ventricle heart palliation. Comput Methods Biomech Biomed Engin. 2014;17(14):1572–1589. doi:10.1080/10255842.2012.758254

31. Schiavazzi DE, Kung EO, Marsden AL, et al. Hemodynamic effects of left pulmonary artery stenosis after superior cavopulmonary connection: A patient-specific multiscale modeling study. Journal of Thoracic and Cardiovascular Surgery. 2015;149(3):689–696.e3. doi:10.1016/j.jtcvs.2014.12.040

32. Yang W, Feinstein JA, Marsden AL. Constrained optimization of an idealized Y-shaped baffle for the Fontan surgery at rest and exercise. Comput Methods Appl Mech Eng. 2010;199(33-36):2135–2149. doi:10.1016/j.cma.2010.03.012

33. Hsia TY, Figliola R, Bove E, et al. Multiscale modelling of single-ventricle hearts for clinical decision support: A leducq transatlantic network of excellence. European Journal of Cardio-thoracic Surgery. 2016;49(2):365–368. doi:10.1093/ejcts/ezv368

34. Pekkan K, Zélicourt D De, Ge L, et al. Physics-driven CFD modeling of complex anatomical cardiovascular flows - A TCPC case study. Ann Biomed Eng. 2005;33(3):284–300. doi:10.1007/s10439-005-1731-0

35. Mynard JP, M.R. Davidson, D.J. Penny, Smolich JJ. A simple, versatile valve model for use in lumped parameter and one-dimensional cardiovascular models. Int J Numer Method Biomed Eng. 2012;28:626–641.

36. Pant S, Corsini C, Baker C, Hsia TY, Pennati G, Vignon-Clementel IE. Data assimilation and modelling of patient-specific single-ventricle physiology with and without valve regurgitation. J Biomech. 2016;49(11):2162–2173. doi:10.1016/j.jbiomech.2015.11.030

37. Yuki K, Emani S, Dinardo JA. A mathematical model of transitional circulation toward biventricular repair in hypoplastic left heart syndrome. Anesth Analg. 2012;115(3):618–626. doi:10.1213/ANE.0b013e31825d36a1

38. Tran JS, Schiavazzi DE, Ramachandra AB, Kahn AM, Marsden AL. Automated tuning for parameter identification and uncertainty quantification in multi-scale coronary simulations. Comput Fluids. 2015;0:1–11. doi:10.1016/j.compfluid.2016.05.015

39. Corsini C, Baker C, Kung E, et al. An integrated approach to patient-specific predictive modeling for single ventricle heart palliation. Comput Methods Biomech Biomed Engin. 2014;17(14):1572–1589. doi:10.1080/10255842.2012.758254

40. Snyder MF, Rideout VC. Computer Simulation Studies of the Venous Circulation. IEEE Trans Biomed Eng. 1969;BME-16(4):325-334. doi:10.1109/TBME.1969.4502663

41. Pennati G, Fumero R. Scaling approach to study the changes through the gestation of human fetal cardiac and circulatory behaviors. Ann Biomed Eng. 2000;28(4):442–452. doi:10.1114/1.282

42. Snyder MF, Rideout VC. Computer Simulation Studies of the Venous Circulation. IEEE Trans Biomed Eng. 1969;(4):325–334. doi:10.1109/TBME.1969.4502663

43. Pennati G, Fumero R. Scaling approach to study the changes through the gestation of human fetal cardiac and circulatory behaviors. Ann Biomed Eng. 2000;28(4):442–452.

44. Tran JS, Schiavazzi DE, Ramachandra AB, Kahn AM, Marsden AL. Automated tuning for parameter identification and uncertainty quantification in multi-scale coronary simulations. Comput Fluids. 2017;142:128–138. doi:10.1016/j.compfluid.2016.05.015

45. Schiavazzi DE, Baretta A, Pennati G, Hsia TY, Marsden AL. Patient-specific parameter estimation in single-ventricle lumped circulation models under uncertainty. Int J Numer Method Biomed Eng. 2017;33(3):1–34. doi:10.1002/cnm.2799

46. Nelder JA, Mead R. A Simplex Method for Function Minimization. Comput J. 1965;7(4). doi:10.1093/comjnl/7.4.308

47. Delmo Walter EM, Ewert P, Hetzer R, et al. Biventricular repair in children with complete atrioventricular septal defect and a small left ventricle. European Journal of Cardio-thoracic Surgery. 2008;33(1):40–47. doi:10.1016/j.ejcts.2007.09.037

48. Conover T, Hlavacek AM, Migliavacca F, et al. An interactive simulation tool for patient-specific clinical decision support in single-ventricle physiology. Journal of Thoracic and Cardiovascular Surgery. 2018;155(2):712–721. doi:10.1016/j.jtcvs.2017.09.046

49. Broomé M, Maksuti E, Bjällmark A, Frenckner B, Janerot-Sjöberg B. Closed-loop real-time simulation model of hemodynamics and oxygen transport in the cardiovascular system. Biomed Eng Online. 2013;12(1):1–20. doi:10.1186/1475-925X-12-69

50. Burkhoff D, Mirsky I, Suga H. Assessment of systolic and diastolic ventricular properties via pressure-volume analysis: A guide for clinical, translational, and basic researchers. Am J Physiol Heart Circ Physiol. 2005;289(2 58-2). doi:10.1152/ajpheart.00138.2005

51. Arts T, Delhaas T, Bovendeerd P, Verbeek X, Prinzen FW. Adaptation to mechanical load determines shape and properties of heart and circulation: The CircAdapt model. Am J Physiol Heart Circ Physiol. 2005;288(4 57-4):1943-1954. doi:10.1152/ajpheart.00444.2004

52. Marsden AL, Feinstein JA. Computational modeling and engineering in pediatric and congenital heart disease. Curr Opin Pediatr. 2015;27(5):587–596.

53. Mittal R, Seo JH, Vedula V, et al. Computational modeling of cardiac hemodynamics: Current status and future outlook. J Comput Phys. 2015;305:1065–1082. doi:10.1016/j.jcp.2015.11.022

54. Haghebaert M, Varsos P, Meiburg R, Vignon-Clementel I. A comparative study of lumped heart models for personalized medicine through sensitivity and identifiability analysis. Journal of Physiology. Published online 2025. doi:10.1113/JP287929

55. Donker DW, Brodie D, Henriques JPS, Broomé M. Left ventricular unloading during veno-arterial ECMO: A simulation study. ASAIO Journal. 2019;65(1):11–20. doi:10.1097/MAT.0000000000000755

56. Yang W, Marsden AL, Ogawa MT, et al. Right ventricular stroke work correlates with outcomes in pediatric pulmonary arterial hypertension. Pulm Circ. 2018;8(3):1–9. doi:10.1177/2045894018780534

57. Sahni A, Marshall L, Cetatoiu MA, et al. Biomechanical Analysis of Age-Dependent Changes in Fontan Power Loss. Ann Biomed Eng. 2024;52(9):2440–2456. doi:10.1007/s10439-024-03534-9

58. Mercer-Rosa L, Fogel MA, Wei ZA, et al. Fontan Geometry and Hemodynamics Are Associated With Quality of Life in Adolescents and Young Adults. Annals of Thoracic Surgery. 2022;114(3):841–847. doi:10.1016/j.athoracsur.2022.01.017

59. Restrepo M, Mirabella L, Tang E, et al. Fontan pathway growth: A quantitative evaluation of lateral tunnel and extracardiac cavopulmonary connections using serial cardiac magnetic resonance. Annals of Thoracic Surgery. 2014;97(3):916–922. doi:10.1016/j.athoracsur.2013.11.015

60. Vedula V, George R, Younes L, Mittal R. Hemodynamics in the left atrium and its effect on ventricular flow patterns. J Biomech Eng. 2015;137:1–8. doi:10.1115/1.4031487

61. Vedula V, Seo JH, Lardo AC, Mittal R. Effect of trabeculae and papillary muscles on the hemodynamics of the left ventricle. Theor Comput Fluid Dyn. 2016;30(1-2):3–21. doi:10.1007/s00162-015-0349-6

62. Seo JH, Vedula V, Abraham T, Mittal R. Multiphysics computational models for cardiac flow and virtual cardiography. Int J Numer Method Biomed Eng. 2013;29:850–869. doi:10.1002/cnm.2556

63. Shi L, Chen I, Takayama H, Vedula V. An Optimization Framework to Personalize Passive Cardiac Mechanics. Comput Methods Appl Mech Eng. 2024;432:117401. doi:10.1016/j.cma.2024.117401

64. Brown AL, Salvador M, Shi L, et al. A modular framework for implicit 3D--0D coupling in cardiac mechanics. Comput Methods Appl Mech Eng. 2024;421:116764.

65. Bäumler K, Vedula V, Sailer AM, et al. Fluid–structure interaction simulations of patient-specific aortic dissection. Biomech Model Mechanobiol. 2020;19(5):1607–1628. doi:10.1007/s10237-020-01294-8

66. Shi L, Chen IY, Vedula V. Personalized Multiscale Modeling of Left Atrial Mechanics and Blood Flow. bioRxiv preprint. Published online 2025. doi:10.1101/2025.04.26.650771

67. Schiavazzi DE, Baretta A, Pennati G, Hsia TY, Marsden AL. Patient-specific parameter estimation in single-ventricle lumped circulation models under uncertainty. Int J Numer Method Biomed Eng. 2017;33(3):1–34. doi:10.1002/cnm.2799

68. Hiebing AA, Pieper RG, Witzenburg CM. A Computational Model of Ventricular Dimensions and Hemodynamics in Growing Infants. J Biomech Eng. 2023;145(10):1–12. doi:10.1115/1.4062779

