## Supplementary Material for "A Patient-specific Computational Model for Neonates and Infants with Borderline Left Ventricles"

### A. List of abbreviations used in the Supplementary Material

| | |
|---|---|
| AOP | Aortic pressure |
| ASD | Atrial septal defect |
| ASDPG | Pressure gradient across the atrial septal defect |
| AVPG | Aortic valve pressure gradient |
| BiVR | Biventricular repair |
| BLV | Borderline left ventricle |
| BSA | Body Surface Area |
| $CV_{O2}$ | Whole body $O_2$ consumption |
| CVP | Central venous pressure |
| EF | Ejection Fraction |
| LAP | Left atrial pressure |
| LPN | Lumped parameter network |
| LVOT | Left ventricular outflow tract |
| MVPG | Pressure gradient across the mitral valve |
| $O_2Cap$ | Maximal $O_2$ carrying capacity |
| $O_2D$ | Oxygen delivery |
| PAP | Pulmonary arterial pressure |
| PDAPG | Pressure gradient across the patent ductus arteriosus |
| PVPG | Pulmonary valve pressure gradient |
| PVR | Pulmonary vascular resistance |
| PVRi | Indexed PVR |
| $Q_{AoV}$ | Aortic valve flow rate |
| Qp | Pulmonary flow rate |
| $Q_{PuV}$ | Pulmonary valve flow rate |
| Qs | Systemic flow rate |
| RAP | Right atrial pressure |
| RVP | Right ventricular pressure |
| RVOT | Right ventricular outflow tract |
| $S_{art}$ | Arterial saturation |
| Std | Standard deviation |
| SV | Stroke volume |
| $S_{ven}$ | Venous saturation |
| S1P | Stage 1 palliation |
| SVR | Systemic vascular resistance |
| SVRi | Indexed SVR |
| VSDPG | Pressure gradient across the ventricular septal defect |
| VTI | Velocity time integral |

### B. Equations governing LPN blocks

Our comprehensive multi-block LPN model to simulate a BLV patient's circulatory system is governed by Kirchhoff's voltage and current laws from circuit theory, mathematically translating to a set of 35 coupled differential-algebraic equations (DAEs) with 89 parameters. These are detailed in the following subsections.

**B1. Heart model**

The time-dependent pressure in each heart chamber ($i$ = LA, RA, LV, RV) is derived from an active and a passive component as[1],

$$P_i(t) = P_{i,active}(t) + P_{i,passive}(t) \tag{1}$$

A time-varying elastance function ($E_i(V_i,\ t)$) governs the active pressure,

$$P_{i,active}(t) \ = \ E_i(V_i,\ t) \ \cdot \ \left(V_i(t) \ - \ V_{0,\,i}\right) \ + \ R_{myo} \cdot \ \frac{dV_i(t)}{dt} \tag{2}$$

where, $V_{0,\,i}$ is the unstressed chamber volume, and the myocardial resistance term ($R_{myo}$) is applied only to ventricles to model viscous effects[1]. The time-varying elastance function is regulated by a sinusoidal function, $A_i(t)$, representing the excitation-relaxation of myocardial contractile elements and a volume-dependent elastance, $E_i^*$,[1]

$$E_i(V_i, t) = A_i(t) \cdot E_i^*(V_i) \tag{3.1}$$

$$E_i^*(V_i) = a_i \cdot \left(V_i - V_{0,i}\right) + b_i \tag{3.2}$$

A non-zero $a_i$ is used only for ventricles, whereas $a_i = 0$ for the atria[1]. The passive pressure follows an exponential relationship,

$$P_{i,passive}(t) = c_i \cdot \{e^{d_i[V_i(t) - V_{0,i}]} - 1\} \tag{4}$$

where $c_i$ and $d_i$ are chamber-specific constant parameters.

**B2. Valve model**

We adopted a nonlinear valve model with a Bernoulli-type resistance ($B$) that governs pressure drop due to convective acceleration and dynamic pressure losses caused by the diverging flow field downstream of the vena contracta. An inertance factor ($L$) controls the pressure drop due to acceleration. The total transvalvular pressure drop is expressed as[2,3],

$$\Delta p = Bq|q| + L\frac{dq}{dt} \tag{5.1}$$

$$B = \frac{\rho}{2A_{eff}^2}, L = \frac{\rho l_{eff}}{A_{eff}} \tag{5.2}$$

where, $l_{eff}$ is the effective length of the valve and $A_{eff}$ is the effective area dependent on the state of the valve. We define an index for the valve state, $\xi$, where $0 \leq \xi \leq 1$ such that $\xi = 0$ indicates a closed valve while $\xi = 1$ indicates an open state. The effective area is then computed as,

$$A_{eff}(t) = A_{eff,max} \cdot \xi(t) + A_{eff,min} \cdot \big(1 - \xi(t)\big) \tag{6}$$

where $A_{eff,min}$ and $A_{eff,max}$ are the minimum and maximum effective areas controlled by the regurgitation ($M_{rg}$) and stenosis ($M_{st}$) factors, respectively,

$$A_{eff,min} = M_{rg}A_{ann};\ A_{eff,max} = M_{st}A_{ann} \tag{7}$$

and $A_{ann}$ is the valve annular area assumed to be fixed. The valve state is determined using rate equations that control the rate at which the valve opens or closes depending on the transvalvular pressure gradients,

$$\frac{d\xi}{dt} = (1 - \xi) \cdot K_{vo} \cdot \Delta p, \qquad \frac{d\xi}{dt} = \xi \cdot K_{vc} \cdot \Delta p \tag{8}$$

where $K_{vo}$ and $K_{vc}$ are the opening and closing rate coefficients, respectively[2,3].

**B3. Transport across the heart chambers**

Mass conservation is applied to transport blood through the heart chamber, where the rate of change of chamber volume is balanced by the net flow rate across it (i.e., net flow = total inflow – total outflow). These are expressed as,

$$\frac{dV_{RA}}{dt} = Q_{SVC} + Q_{THIVC} + Q_{ASD} - Q_{TV} \tag{9.1}$$

$$\frac{dV_{LA}}{dt} = Q_{RPV} + Q_{LPV} - Q_{ASD} - Q_{MV} \tag{9.2}$$

$$\frac{dV_{RV}}{dt} = Q_{TV} + Q_{VSD} - Q_{PulV} \tag{9.3}$$

$$\frac{dV_{LV}}{dt} = Q_{MV} - Q_{VSD} - Q_{AoV} \tag{9.4}$$

In the above equations, $Q_{ASD}, Q_{VSD}$ are the flow rates through the atrial and ventricular septal defects, respectively, while $Q_{TV}, Q_{MV}, Q_{PulV}, Q_{AoV}$ are the flow rates through the tricuspid, mitral, pulmonary, and aortic valves, respectively, governed by the adopted valve model (Sec. B2) as,

$$L_{TV}\frac{dQ_{TV}}{dt} = P_{RA} - P_{RV} - B_{TV}Q_{TV}|Q_{TV}| \tag{10.1}$$

$$L_{MV}\frac{dQ_{TV}}{dt} = P_{LA} - P_{LV} - B_{MV}Q_{MV}|Q_{MV}| \tag{10.2}$$

$$L_{PulV}\frac{dQ_{PulV}}{dt} = P_{RV} - P_{PA} - B_{PulV}Q_{PulV}|Q_{PulV}| \tag{10.3}$$

$$L_{AoV}\frac{dQ_{AoV}}{dt} = P_{LV} - P_{Ao} - B_{AoV}Q_{AoV}|Q_{AoV}| \tag{10.4}$$

**B4. Systemic and pulmonary circulation**

Outside the heart compartment, the systemic and pulmonary circulation blocks are generally modeled using R-L-C Windkessel circuit elements describing the circulation following

Kirchhoff's voltage and current laws at each node. For any generic block, $i \in$ $ThAo, AbAo, Li, I, K, LEGA, LEGV,\ AbIVC, ThIVC, UB, SVC$,

$$C_i \frac{dP_i}{dt} = Q_{u,i} - Q_{d,i}, \qquad L_i \frac{dQ_i}{dt} = P_{u,i} - P_{d,i} - R_i Q_i \tag{11}$$

where $Q_u$ and $Q_d$ are the upstream and downstream flows into and out of the node, $P_u$ and $P_d$ are the upstream and downstream pressures across the inductor-resistor element[4].

**B5. Blalock-Taussig-Thomas shunt for Norwood (S1P)**

A nonlinear convective energy loss function, accounting for the viscous and inertial effects, models the pressure drop across the BTT shunt after S1P surgery.[1]

$$\Delta P = R_{sh} Q_{sh} + K_{sh} Q_{sh}^2 = \frac{k_1}{D^4} Q_{sh} + \frac{k_2}{D^4} Q_{sh}^2 \tag{12}$$

where $Q_{sh}$ is the shunt flow rate and $R_{sh}$ and $K_{sh}$ are the linear and nonlinear shunt resistance parameters, respectively, expressed as a function of the shunt diameter, *D*, with constants $k_1$ and $k_2$ evaluated from prior finite element simulations[1,5].

**B6. Oxygen transport model**

The oxygen transportation model employed here is based on the blood-mixing models proposed by Yuki et al.[6] for the BLV patients and Migliavacca et al.[1] for S1P circulation. A few additional considerations are discussed below, particularly in determining the blood oxygen content of systemic and pulmonary arteries ($C_{sys\text{-}art}$, $C_{pul\text{-}art}$) that are crucial in determining the amount of oxygen delivered to the body.

1. For the preoperative BLV patients with left-to-right shunted ASDs and VSDs, we assumed no reduction in oxygen content in the left heart. Therefore, we have,

$$C_{LA} = C_{LV} = C_{PV} \tag{13.1}$$

$$C_{RV} \cdot Q_{PuV} = C_{sys-ven} \cdot Q_s + C_{LA} \cdot Q_{ASD} + C_{LV} \cdot Q_{VSD} \tag{13.2}$$

where $C_{(\cdot)}$ denotes oxygen content in the blood (mL/dL blood) in the corresponding chamber and $Q_{ASD}$ and $Q_{VSD}$ represent flow rates (L/min) across the atrial and ventricular septal defects, respectively. Further, these patients typically exhibited bidirectional flow across the PDA connecting the aortic arch and the pulmonary artery. We assumed that the ventricular blood is isolated from the PDA shunted blood so that there is no mixing or retrograde flow from the PDA to the ventricle. Then, the systemic and pulmonary arterial oxygen content ($C_{sys\text{-}art}$, $C_{pul\text{-}art}$) relations are given by,

$$C_{sys-art} \cdot Q_s = C_{RV} \cdot Q_{PDA-R} + C_{LV} \cdot Q_{AoV} \tag{14.1}$$

$$C_{pul-art} \cdot Q_p = C_{LV} \cdot Q_{PDA-L} + C_{RV} \cdot Q_{PulV} \tag{14.2}$$

where $Q_{PDA\text{-}R}$ is the flow rate in the PDA shunt from the pulmonary artery to the aorta, $Q_{PDA\text{-}L}$ is the retrograde flow in the PDA shunt from the aorta to the pulmonary artery, $Q_{AoV}$ is the flow rate across the aortic valve, and $Q_{PulV}$ is the flow rate across the pulmonary valve. Finally, we have the relation for the whole-body oxygen consumption ($C\dot{V}_{O2}$) as,

$$(C_{sys-art} - C_{sys-ven}) \cdot Q_s = C\dot{V}_{O2} \tag{15}$$

We assumed $C\dot{V}_{O2}$ as 170 mL/min/m$^2$ for patients who did not undergo cath. The oxygen content in the blood is related to the oxygen saturation by[1],

$$C_{(\cdot)} = Sat_{(\cdot)} \cdot O_2Cap \tag{16}$$

where $O_2Cap = 1.34 \cdot Hb$, is the maximal oxygen-carrying capacity of the blood, where $Hb$ is the hemoglobin content of patients. To close the above system of equations (Eqs. 13–16), we assume that the oxygen saturation in the pulmonary veins ($Sat_{PV}$) is 98%, which is reasonable in the absence of any pulmonary venous disease[5]. Thus, equation set (Eqs. 13–16) is solved to determine the systemic arterial ($S_{art}$ or $Sat_{sys-art}$) and the systemic venous ($S_{ven}$ or $Sat_{sys-ven}$) saturations. The oxygen delivery ($O_2D$) is then calculated as,

$$O_2D = C_{sys-art} \cdot Q_s \tag{17}$$

2. For patients undergoing BiVR, we closed all septal defects and assumed no blood mixing in the oxygen transport model. Therefore, we have,

$$C_{sys-art} = C_{PV} = 0.98\ O_2Cap \tag{18}$$

The equation set (Eqs. 15–17) is used to determine venous saturation and oxygen delivery.

3. For the S1P model governing the Norwood circulation, while the equation set (Eqs. 13–17) is still applied, we account for the BTT shunt based on the blood mixing model proposed by Migliavacca et al.[1].

### C. Method to estimate hemodynamic targets only using echo data

In the absence of cath data, we use Doppler-based pressure gradients and cuff pressures to obtain all the other hemodynamic quantities set as clinical targets for the parameter estimation framework. We obtain $AOP_{min/max/mean}$ from the patients' cuff pressure measurements and assume the central venous pressure to be 5mmHg[7,8].

1. We then use pressure gradients from Doppler waveforms to estimate other hemodynamic targets, including $PAP_{min/max/mean}$ and $LAP_{mean}$ as,[9,10]

$$LVP_{max} = AOP_{max} + AVPG_{max} \tag{19.1}$$

$$RVP_{max} = LVP_{max} - VSDPG_{max} \tag{19.2}$$

$$PAP_{max} = RVP_{max} - PVPG_{max} \tag{19.3}$$

$$PAP_{mean} = AOP_{mean} - PDAPG_{mean} \tag{19.4}$$

$$PAP_{min} = 0.5(3 * PAP_{mean} - PAP_{max}) \tag{19.5}$$

$$LAP_{mean} = RAP_{mean} - ASD_{grad} \tag{19.6}$$

2. We extract the systemic ($Q_s$) and pulmonary ($Q_p$) flow rates using velocity time integrals (VTI) of the Doppler waveforms across LVOT, RVOT, and PDA as,

$$Qs = A_{LVOT} \cdot LVOT_{VTI} - A_{PDA} \cdot PDA_{VTI} \tag{20.1}$$

$$Qp = A_{RVOT} \cdot RVOT_{VTI} + A_{PDA} \cdot PDA_{VTI} \tag{20.2}$$

3. Finally, we estimate the vascular resistances $SVR$ and $PVR$ as $SVR = (AOP_{mean} - RAP_{mean})/Q_s$, and $PVR = (PAP_{mean} - LAP_{mean})/Q_p$, where the mean right atrial pressure ($RAP_{mean}$) is assumed to be the same as the central venous pressure ($RAP_{mean}$ = $CVP$ = 5mmHg).

### SUPPLEMENTARY REFERENCES


1. Migliavacca F, Pennati G, Dubini G, et al. Modeling of the Norwood circulation: Effects of shunt size, vascular resistances, and heart rate. *Am J Physiol Heart Circ Physiol*. 2001;280(5 49-5):2076-2086. doi:10.1152/ajpheart.2001.280.5.h2076

2. Mynard JP, M.R.Davidson, D.J.Penny, Smolich JJ. A simple, versatile valve model for use in lumped parameter and one-dimensional cardiovascular models. *Int J Numer Method Biomed Eng*. 2012;28:626-641.

3. Pant S, Corsini C, Baker C, Hsia TY, Pennati G, Vignon-Clementel IE. Data assimilation and modelling of patient-specific single-ventricle physiology with and without valve regurgitation. *J Biomech*. 2016;49(11):2162-2173. doi:10.1016/j.jbiomech.2015.11.030

4. Corsini C, Baker C, Kung E, et al. An integrated approach to patient-specific predictive modeling for single ventricle heart palliation. *Comput Methods Biomech Biomed Engin*. 2014;17(14):1572-1589. doi:10.1080/10255842.2012.758254

5. Migliavacca F, Dubini G, Pennati G, et al. Computational model of the fluid dynamics in systemic-to-pulmonary shunts. *J Biomech*. 2000;33(5):549-557. doi:10.1016/S0021-9290(99)00219-5

6. Yuki K, Emani S, Dinardo JA. A mathematical model of transitional circulation toward biventricular repair in hypoplastic left heart syndrome. *Anesth Analg*. 2012;115(3):618-626. doi:10.1213/ANE.0b013e31825d36a1

7. Burnard ED, James LS. Atrial pressures and cardiac size in the newborn infant. *J Pediatr*. 1963;62(6):815-826. doi:10.1016/s0022-3476(63)80095-5

8. Skinner JR, Milligan DWA, Hunter S, Hey EN. Central venous pressure in the ventilated neonate. *Arch Dis Child*. 1992;67(4 SPEC NO):374-377. doi:10.1136/adc.67.4_Spec_No.374

9. Harris P, Kuppurao L. Quantitative Doppler echocardiography. *BJA Educ*. 2016;16(2):46-52. doi:10.1093/bjaceaccp/mkv015

10. Parasuraman S, Walker S, Loudon BL, et al. Assessment of pulmonary artery pressure by echocardiography-A comprehensive review. *IJC Heart and Vasculature*. 2016;12:45-51. doi:10.1016/j.ijcha.2016.05.011

### SUPPLEMENTARY TABLES LEGEND

**Supplementary Table ST1.** Clinical data for the BLV patients included in the study.

**Supplementary Table ST2.** Preoperative clinical measurements, set as hemodynamic targets for the LPN parameter estimation algorithm, for the BLV patients in Group I. The corresponding standard deviations (STD) and weights are also included.

**Supplementary Table ST3.** Preoperative clinical measurements, set as hemodynamic targets for the LPN parameter estimation algorithm, for the BLV patients in Group II. The corresponding standard deviations (STD) and weights are also included.

**Supplementary Table ST4.** Preoperative LPN model parameters for the BLV patients in Group I. The parameters are highlighted based on whether they are directly incorporated from clinical data, estimated from scaling analysis, or tuned using the automatic tuning framework.

**Supplementary Table ST5.** Preoperative LPN model parameters for the BLV patients in Group II. The parameters are highlighted based on whether they are directly incorporated from clinical data, estimated from scaling analysis, or tuned using the automatic tuning framework.

**Supplementary Table ST6.** Hemodynamic quantities predicted by the preoperative LPN model for the patients in Group I. The error in LPN prediction, measured as a (%) of the clinical data, is shown in parentheses.

**Supplementary Table ST7.** Hemodynamic quantities predicted by the preoperative LPN model for the patients in Group II. The error in LPN prediction, measured as a (%) of the clinical data, is shown in parentheses.

**Supplementary Table ST8.** Predicted hemodynamic variables following virtual BiVR and S1P surgical procedures for the patients in Group I.

**Supplementary Table ST9.** Predicted hemodynamic variables following virtual BiVR and S1P surgical procedures for the patients in Group II.

SUPPLEMENTAL TABLES

**SUPPLEMENTARY TABLE ST1. Clinical data for the BLV patients included in the study.**

| | Group I (S1P) | | | | | Group II (BiVR) | | | | |
|---|---|---|---|---|---|---|---|---|---|---|
| **Patient / Clinical Attribute** | **A*** | **B*** | **C** | **D** | **E** | **F** | **G** | **H†** | **I†** | **J†** |
| **Age (months)** | 3 | 2 | 1 | 2 | 2 | 4 | 1 | <1 | <1 | <1 |
| **Weight (kg)** | 3.4 | 5.2 | 3.6 | 2.2 | 8.3 | 4.6 | 4.1 | 3.8 | 2.9 | 3.1 |
| **BSA ($m^2$)** | 0.29 | 0.26 | 0.27 | 0.21 | 0.21 | 0.25 | 0.23 | 0.24 | 0.21 | 0.18 |
| **MV Z-score** | -4.53 | -5.80 | -4.20 | -5.00 | 0.30 | -1.30 | -1.97 | -2.86 | -1.90 | -0.20 |
| **AV Z-score** | -5.58 | -5.73 | -2.69 | -3.47 | -1.77 | -0.22 | -4.05 | -4.62 | -1.14 | -2.22 |
| **Mechanical Ventilation** | No | No | Yes | Yes | No | No | Yes | No | No | No |
| **Inotropes Requirement** | No | No | Yes | No | No | No | No | No | No | No |
| **Hypoplastic Aortic Arch/Coarctation of Aorta** | Yes | Yes | Yes | Yes | Yes | No | No | Yes | Yes | Yes |
| **ASD** | Yes | Yes | Yes | Yes | Yes | Yes | Yes | Yes | Yes | Yes |
| **VSD** | No | Yes | Yes | Yes | No | Yes | No | Yes | Yes | No |
| **Mitral Stenosis** | Yes | Yes | Yes | Yes | No | Yes | No | Yes | No | No |
| **Aortic Stenosis** | Yes | Yes | No | No | Yes | Yes | Yes | Yes | No | No |
| **$CV_{O2}$ (cath) (mL/min/$m^2$)** | 170 | 170 | 180 | 160 | 150 | 170 | 150 | - | - | - |
| **LVEDV (mL)** | 4.0 | 8.0 | 7.1 | - | 7.1 | - | 6.7 | 4.8 | 6.4 | 6.9 |
| **LVEDV Z-score** | -4.89 | -1.09 | -1.45 | - | -1.01 | - | -2.20 | -3.96 | -1.68 | 0.35 |
| **Qp/Qs** | 0.98 | 1.07 | 1.93 | 2.97 | 2.00 | 3.78 | 3.40 | 3.23 | 3.53 | 3.26 |
| **$PAP_{mean}$ (mmHg)** | 44.00 | 61.00 | 41.00 | 35.00 | 35.00 | 22.00 | 28.00 | 39.00 | 19.00 | 33.00 |

| | | | | | | | | | | |
|---|---|---|---|---|---|---|---|---|---|---|
| **$AOP_{mean}$ (mmHg)** | 49.00 | 60.00 | 53.00 | 36.00 | 44.00 | 54.00 | 56.00 | 50.00 | 46.00 | 55.00 |
| **$LAP_{mean}$ (mmHg)** | 5.00 | 5.00 | 23.00 | 14.00 | 11.00 | 5.00 | 11.00 | 13.00 | 5.00 | 13.00 |
| **PVRi (WU • $m^2$)** | 2.50 | 3.44 | 4.10 | 3.10 | 2.40 | 1.90 | 2.47 | 3.49 | 1.69 | 3.12 |
| **SVRi (WU • $m^2$)** | 11.60 | 13.82 | 12.60 | 9.90 | 5.90 | 19.60 | 20.96 | 27.92 | 17.90 | 21.92 |

*patients underwent pulmonary artery banding; †data collected based on echocardiography alone;

**SUPPLEMENTARY TABLE ST2. Preoperative clinical measurements, set as hemodynamic targets for the LPN parameter estimation algorithm, for the BLV patients in Group I. The corresponding standard deviations (STD) and weights are also included.**

| Patient / Clinical Target | A† | B† | C | D | E | STD | weight |
|---|---|---|---|---|---|---|---|
| $AOP_{min}$ | 41.00 | 37.00 | 34.00 | 28.00 | 33.00 | ±10% | 1 |
| $AOP_{max}$ | 65.00 | 94.00 | 96.00 | 52.00 | 67.00 | ±10% | 1 |
| $AOP_{mean}$ | 49.00 | 60.00 | 53.00 | 36.00 | 44.00 | ±10% | 1 |
| $PAP_{min}$ | 34.00 | 36.00 | 31.00 | 27.00 | 27.00 | ±20% | 1 |
| $PAP_{max}$ | 60.00 | 108.00 | 53.00 | 40.00 | 49.00 | ±20% | 1 |
| $PAP_{mean}$ | 44.00 | 61.00 | 41.00 | 35.00 | 35.00 | ±20% | 1 |
| $LAP_{mean}$ | 5.00 | 5.00 | 23.00 | 14.00 | 11.00 | ±30% | 1 |
| **Qp/Qs** | 0.98 | 1.07 | 1.93 | 2.97 | 2.00 | ±20% | 1 |
| **PVRi** | 2.50 | 3.44 | 4.10 | 3.10 | 2.40 | ±10% | 1 |
| **SVRi** | 11.60 | 13.82 | 12.60 | 9.90 | 5.90 | ±10% | 1 |
| **SV (mL)** | - | - | - | - | - | ±20% | 2 |
| **EF (%)** | - | - | - | - | - | ±20% | 2 |
| $MVPG_{max}$ | 6.00 | 9..00 | 17.00 | 8.00 | 11.00 | ±20% | 2 |
| $AVPG_{max}$ | - | - | - | - | 26.00 | ±20% | 2 |
| $PVPG_{max}$ | - | 8.00 | - | - | - | ±20% | 2 |
| $VSDPG_{mean}$ | - | - | - | - | - | ±20% | 2 |
| $ASDPG_{mean}$ | 1.00 | 1.00 | 10.00 | 6.00 | 2.50 | ±20% | 2 |
| $PDAPG_{mean}$ | 5.00 | - | 12.00 | 1.00 | 9.00 | ±20% | 2 |

†patients underwent pulmonary artery banding; All pressures and pressure gradients are in *mmHg*; PVRi/SVRi: $WU \cdot m^2$.

**SUPPLEMENTARY TABLE ST3. Preoperative clinical measurements, set as hemodynamic targets for the LPN parameter estimation algorithm, for the BLV patients in Group II. The corresponding standard deviations (STD) and weights are also included.**

| Patient / Clinical Target | F | G | H† | I† | J† | STD | weight |
|---|---|---|---|---|---|---|---|
| $AOP_{min}$ | 42.00 | 45.00 | 41.00 | 39.00 | 48.00 | ±10% | 1 |
| $AOP_{max}$ | 75.00 | 66.00 | 68.00 | 60.00 | 67.00 | ±10% | 1 |
| $AOP_{mean}$ | 54.00 | 56.00 | 50.00 | 46.00 | 55.00 | ±10% | 1 |
| $PAP_{min}$ | 16.00 | 23.00 | 32.00 | 15.00 | 29.00 | ±20% | 1 |
| $PAP_{max}$ | 30.00 | 35.00 | 47.00 | 27.00 | 37.00 | ±20% | 1 |
| $PAP_{mean}$ | 22.00 | 28.00 | 39.00 | 19.00 | 33.00 | ±20% | 1 |
| $LAP_{mean}$ | 5.00 | 11.00 | 13.00 | 5.00 | 13.00 | ±30% | 1 |
| Qp/Qs | 3.78 | 3.40 | 3.23 | 3.53 | 3.26 | ±20% | 1 |
| PVRi | 1.90 | 2.47 | 3.49 | 1.69 | 3.12 | ±10% | 1 |
| SVRi | 19.60 | 20.96 | 27.92 | 17.90 | 21.92 | ±10% | 1 |
| SV (mL) | - | - | 3.20 | 3.20 | 2.80 | ±20% | 2 |
| EF (%) | - | 58.00 | 64.00 | 50.00 | 55.00 | ±20% | 2 |
| $MVPG_{max}$ | 8.00 | - | 6.00 | 8.50 | 10.00 | ±20% | 2 |
| $AVPG_{max}$ | 11.00 | - | 13.00 | 2.00 | 33.00 | ±20% | 2 |
| $PVPG_{max}$ | - | 6.00 | 8.00 | 12.00 | 2.00 | ±20% | 2 |
| $VSDPG_{mean}$ | - | - | 23.00 | 22.00 | - | ±20% | 2 |
| $ASDPG_{mean}$ | 2.00 | 8.00 | 9.00 | 1.00 | 12.00 | ±20% | 2 |
| $PDAPG_{mean}$ | 32.00 | 22.00 | 11.00 | - | 22.00 | ±20% | 2 |

†data collected based on echocardiography alone; All pressures and pressure gradients are in *mmHg*; PVRi/SVRi: $WU \cdot m^2$.

**SUPPLEMENTARY TABLE ST4. Preoperative LPN model parameters for the BLV patients in Group I. The parameters are highlighted based on whether they are directly incorporated from clinical data, estimated from scaling analysis, or tuned using the automatic tuning framework.**

| | Fixed clinical inputs | | Automatic tuning framework | | Parameters from scaling |
|---|---|---|---|---|---|

| Patient / LPN parameter | A† | B† | C | D | E |
|---|---|---|---|---|---|
| $\boldsymbol{T}$ | 0.4620 | 0.4000 | 0.4290 | 0.4000 | 0.4050 |
| $\boldsymbol{b_A}$ | 1.5030 | 1.5293 | 1.9492 | 1.0230 | 1.5020 |
| $\boldsymbol{c_A}$ | 0.4100 | 0.4100 | 0.4000 | 0.2040 | 0.4100 |
| $\boldsymbol{d_A}$ | 0.4000 | 0.4000 | 0.4000 | 0.2190 | 0.4000 |
| $\boldsymbol{V_{0,A}}$ | 1.6300 | 1.6300 | 0.9000 | 0.6710 | 1.6300 |
| $\boldsymbol{a_{RV}}$ | -0.1150 | -0.1150 | -0.1120 | -0.1120 | -0.1150 |
| $\boldsymbol{b_{RV}}$ | 11.0000 | 12.0000 | 18.0000 | 6.0200 | 12.0000 |
| $\boldsymbol{c_{RV}}$ | 0.2500 | 0.2500 | 0.2600 | 0.1280 | 0.2500 |
| $\boldsymbol{d_{RV}}$ | 0.1400 | 0.1400 | 0.2100 | 0.1080 | 0.1700 |
| $\boldsymbol{V_{0,RV}}$ | 4.1000 | 4.2000 | 2.9000 | 4.9600 | 2.8000 |
| $\boldsymbol{a_{LV}}$ | -0.0800 | -0.0840 | -0.0840 | -0.0160 | -0.0840 |
| $\boldsymbol{b_{LV}}$ | 30.0000 | 45.0000 | 32.0000 | 51.6000 | 60.0000 |
| $\boldsymbol{c_{LV}}$ | 0.9000 | 0.9000 | 0.4500 | 0.2160 | 0.6000 |
| $\boldsymbol{d_{LV}}$ | 0.4200 | 0.4200 | 0.3800 | 0.4490 | 0.3900 |
| $\boldsymbol{V_{0,LV}}$ | 1.2000 | 1.2000 | 0.9000 | 2.1000 | 0.1600 |

| | | | | | |
|---|---|---|---|---|---|
| $R_{ASD}$ | 0.1000 | 0.1000 | 1.0000 | 0.3090 | 0.1000 |
| $R_{VSD}$ | $10^5$ | $10^5$ | 1.0000 | $10^5$ | $10^5$ |
| $K_{TVst}$ | 1.0000 | 1.0000 | 1.0000 | 1.0000 | 1.0000 |
| $K_{TVrg}$ | 0.0000 | 0.0000 | 0.0000 | 0.0000 | 0.0000 |
| $K_{TVvo}$ | 0.0300 | 0.0300 | 0.0300 | 0.0300 | 0.0300 |
| $K_{TVvc}$ | 0.0400 | 0.0400 | 0.0400 | 0.0400 | 0.0400 |
| $K_{MVst}$ | 0.5000 | 0.5000 | 0.5000 | 0.5000 | 1.0000 |
| $K_{MVrg}$ | 0.0000 | 0.0000 | 0.0000 | 0.0000 | 0.0000 |
| $K_{MVvo}$ | 0.0300 | 0.0300 | 0.0300 | 0.0300 | 0.0300 |
| $K_{MVvc}$ | 0.0400 | 0.0400 | 0.0400 | 0.0400 | 0.0400 |
| $K_{AVst}$ | 0.3000 | 0.3000 | 0.3000 | 0.5000 | 0.5000 |
| $K_{AVrg}$ | 0.0000 | 0.0000 | 0.0000 | 0.0000 | 0.0000 |
| $K_{AVvo}$ | 0.0900 | 0.0900 | 0.0900 | 0.0900 | 0.0900 |
| $K_{AVvc}$ | 0.0900 | 0.0900 | 0.0900 | 0.0900 | 0.0900 |
| $K_{PVst}$ | 1.0000 | 1.0000 | 1.0000 | 1.0000 | 1.0000 |
| $K_{PVrg}$ | 0.0000 | 0.0000 | 0.0000 | 0.0000 | 0.0000 |
| $K_{PVvo}$ | 0.1800 | 0.1800 | 0.1800 | 0.1800 | 0.1800 |
| $K_{PVvc}$ | 0.1800 | 0.1800 | 0.1800 | 0.1800 | 0.1800 |
| $C_{Ao}$ | 0.1170 | 0.1020 | 0.0791 | 0.0524 | 0.0624 |
| $C_{Arc}$ | 0.1170 | 0.1020 | 0.1020 | 0.0524 | 0.0624 |
| $R_{AoA}$ | 1.3200 | 1.8000 | 1.8000 | 0.1770 | 0.1770 |
| $C_{UB}$ | 1.7000 | 1.8800 | 1.2400 | 1.4700 | 3.2100 |

| | | | | | |
|---|---|---|---|---|---|
| $L_{UBa}$ | 0.0007 | 0.0007 | 0.0007 | 0.0007 | 0.0007 |
| $R_{UBa}$ | 0.8840 | 1.1300 | 1.1200 | 0.9870 | 0.5480 |
| $R_{UBv}$ | 2.5800 | 3.3000 | 3.2600 | 2.8800 | 1.6000 |
| $C_{SVC}$ | 0.1870 | 0.1900 | 0.1060 | 0.1610 | 0.2730 |
| $R_{SVC}$ | 0.1080 | 0.1470 | 0.1650 | 0.1200 | 0.0812 |
| $C_{ThAo}$ | 0.0245 | 0.0248 | 0.0301 | 0.0211 | 0.0358 |
| $L_{ThAo}$ | 0.0023 | 0.0024 | 0.0025 | 0.0022 | 0.0023 |
| $R_{ThAo}$ | 0.2570 | 0.3520 | 0.2200 | 0.2870 | 0.1940 |
| $C_{AbAo}$ | 0.0517 | 0.0525 | 0.0638 | 0.0446 | 0.0756 |
| $L_{AbAo}$ | 0.0023 | 0.0024 | 0.0025 | 0.0022 | 0.0023 |
| $R_{AbAo}$ | 1.9600 | 2.6800 | 1.6800 | 2.1900 | 1.4800 |
| $C_{Lvr}$ | 0.6440 | 0.6530 | 0.7930 | 0.5560 | 0.9410 |
| $R_{Lvra}$ | 30.6000 | 41.8000 | 26.1000 | 34.1000 | 23.0000 |
| $R_{Lvrv}$ | 0.1920 | 0.2620 | 0.1640 | 0.2140 | 0.1440 |
| $C_{Kd}$ | 0.2680 | 0.2720 | 0.3300 | 0.2310 | 0.3910 |
| $R_{Kda}$ | 21.6000 | 29.5000 | 18.4000 | 24.1000 | 16.2000 |
| $R_{Kdv}$ | 2.0200 | 2.7600 | 1.7300 | 2.2500 | 1.5200 |
| $C_{I}$ | 0.1600 | 0.1630 | 0.1980 | 0.1380 | 0.2340 |
| $R_{Ia}$ | 47.3000 | 64.7000 | 40.4000 | 52.8000 | 35.6000 |
| $R_{Iv}$ | 0.8600 | 1.1800 | 0.7350 | 0.9600 | 0.6470 |
| $C_{Lega}$ | 0.0205 | 0.0181 | 0.0167 | 0.0177 | 0.0198 |
| $L_{Lega}$ | 0.0023 | 0.0024 | 0.0025 | 0.0022 | 0.0023 |

| | | | | | |
|---|---|---|---|---|---|
| $R_{Lega}$ | 6.2200 | 9.4500 | 7.2500 | 6.9500 | 6.3800 |
| $R_{Legc}$ | 14.7000 | 22.3000 | 17.1000 | 16.4000 | 15.0000 |
| $C_{Legv}$ | 0.4180 | 0.3690 | 0.3410 | 0.3610 | 0.4040 |
| $R_{Legv}$ | 3.4300 | 5.2100 | 3.9900 | 3.8300 | 3.5100 |
| $C_{AbIV}$ | 0.2660 | 0.2700 | 0.3280 | 0.2300 | 0.3890 |
| $R_{AbIV}$ | 0.0539 | 0.0737 | 0.0461 | 0.0602 | 0.0405 |
| $C_{ThIV}$ | 0.7970 | 0.8090 | 0.9830 | 0.6880 | 1.1700 |
| $R_{ThIV}$ | 0.0420 | 0.0575 | 0.0359 | 0.0469 | 0.0316 |
| $R_{PDA}$ | 0.0100 | 0.1000 | 0.1170 | 9.3300 | 1.1700 |
| $C_{PA}$ | 0.2260 | 0.1990 | 0.1540 | 0.1730 | 0.1540 |
| $C_{PA2}$ | 0.2260 | 0.1990 | 0.1540 | 0.1730 | 0.1540 |
| $R_{T}$ | 2.0000 | 2.0000 | 0.0100 | 0.0020 | 0.0020 |
| $RCRr_{C1}$ | 0.1690 | 0.1660 | 0.0663 | 0.3220 | 0.0363 |
| $RCRr_{C2}$ | 0.0347 | 0.0340 | 0.0136 | 0.0661 | 0.0075 |
| $RCRr_{C3}$ | 1.5700 | 1.5400 | 0.6170 | 3.0000 | 0.3380 |
| $RCRr_{Rd1}$ | 5.0500 | 5.1300 | 10.2000 | 3.1200 | 7.0800 |
| $RCRr_{Rd2}$ | 21.4000 | 21.7000 | 43.2000 | 13.2000 | 29.9000 |
| $RCRr_{Rd3}$ | 0.6820 | 0.6920 | 1.3800 | 0.4210 | 0.9550 |
| $RCRr_{Rp1}$ | 1.0400 | 1.0500 | 2.0900 | 0.6380 | 1.4400 |
| $RCRr_{Rp2}$ | 2.6400 | 2.6800 | 5.3300 | 1.6300 | 3.7000 |
| $RCRr_{Rp3}$ | 0.3360 | 0.3410 | 0.6780 | 0.2070 | 0.4700 |
| $RCRl_{C1}$ | 0.0263 | 0.0258 | 0.0103 | 0.0502 | 0.0057 |

| | | | | | |
|---|---|---|---|---|---|
| $RCRl_{C2}$ | 0.0650 | 0.0637 | 0.0255 | 0.1240 | 0.0140 |
| $RCRl_{C3}$ | 0.8190 | 0.8030 | 0.3210 | 1.5600 | 0.1760 |
| $RCRl_{Rd1}$ | 30.7000 | 31.2000 | 62.0000 | 18.9000 | 43.0000 |
| $RCRl_{Rd2}$ | 13.0000 | 13.2000 | 26.3000 | 8.0300 | 18.2000 |
| $RCRl_{Rd3}$ | 1.6400 | 1.6600 | 3.3000 | 1.0100 | 2.2900 |
| $RCRl_{Rp1}$ | 0.6270 | 0.6360 | 1.2600 | 0.3860 | 0.8770 |
| $RCRl_{Rp2}$ | 0.2660 | 0.2700 | 0.5370 | 0.1640 | 0.3720 |
| $RCRl_{Rp3}$ | 0.0682 | 0.0692 | 0.1380 | 0.0420 | 0.0955 |

**Units:** ***T*** (Cardiac cycle duration, *s*), ***c*** (Elastance factor of the cardiac chambers, *mmHg*), ***d*** (Diastolic exponential factor of the cardiac chambers, $cm^{-3}$), $V_0$ (Unstressed volume, $cm^3$), ***a*** (Quadratic elastance factor for the ventricles, $mmHg.cm^{-6}$), ***b*** (Linear elastance factor for the ventricles, $mmHg.cm^{-3}$), ***R*** (Vascular resistance, $mmHg.s.cm^{-3}$), ***L*** (Vascular inductance/inertia, $mmHg.s^2.cm^{-3}$), $RCR_C$ (Compliance of the pulmonary RCR block, $cm^3.mmHg^{-1}$), $RCR_{Rd}$ (Distal resistance of the pulmonary RCR block, $mmHg.s.cm^{-3}$), $RCR_{Rp}$ (Proximal resistance of the pulmonary RCR block, $mmHg.s.cm^{-3}$); †patients underwent pulmonary artery banding;

| | | | | | |
|---|---|---|---|---|---|
| (grey) | Fixed clinical inputs | (orange) | Automatic tuning framework | (blue) | Parameters from scaling |

**SUPPLEMENTARY TABLE ST5. Preoperative LPN model parameters for the BLV patients in Group II. The parameters are highlighted based on whether they are directly incorporated from clinical data, estimated from scaling analysis, or tuned using the automatic tuning framework.**

| | Fixed clinical inputs | | Automatic tuning framework | | Parameters from scaling |
|---|---|---|---|---|---|

| Patient / LPN parameter | F | G | H† | I† | J† |
|---|---|---|---|---|---|
| $T$ | 0.4620 | 0.3590 | 0.4000 | 0.4000 | 0.4000 |
| $b_A$ | 0.3330 | 0.5000 | 0.7350 | 0.7350 | 0.7350 |
| $c_A$ | 0.2700 | 0.2700 | 0.9060 | 0.7540 | 0.9060 |
| $d_A$ | 0.4000 | 0.4000 | 0.5550 | 0.4550 | 0.5550 |
| $V_{A0}$ | 0.9000 | 0.9000 | 0.5080 | 0.5080 | 0.5080 |
| $a_{RV}$ | -0.0420 | -0.0230 | -0.0120 | -0.0120 | -0.0120 |
| $b_{RV}$ | 35.0000 | 19.0000 | 12.0000 | 5.9100 | 16.0000 |
| $c_{RV}$ | 0.0900 | 0.0900 | 0.1300 | 0.1470 | 0.1300 |
| $d_{RV}$ | 0.0400 | 0.0900 | 0.2400 | 0.1270 | 0.1200 |
| $V_{0,RV}$ | 4.8000 | 4.0000 | 3.9000 | 4.7200 | 3.9000 |
| $a_{LV}$ | -0.0840 | -0.0840 | -0.0120 | -0.0120 | -0.0120 |
| $b_{LV}$ | 40.0000 | 37.0000 | 25.3000 | 12.3000 | 30.3000 |
| $c_{LV}$ | 0.4700 | 0.7600 | 0.4100 | 0.4950 | 0.4100 |
| $d_{LV}$ | 0.3200 | 0.3200 | 0.2500 | 0.1910 | 0.3100 |
| $V_{0,LV}$ | 1.8000 | 2.3000 | 0.6050 | 0.3550 | 0.3050 |

| | | | | | |
|---|---|---|---|---|---|
| $R_{ASD}$ | 0.0100 | 1.0000 | 0.7000 | 0.0358 | 4.0000 |
| $R_{VSD}$ | $10^5$ | $10^5$ | $10^5$ | 1.5000 | $10^5$ |
| $K_{TVst}$ | 1.0000 | 1.0000 | 1.0000 | 1.0000 | 1.0000 |
| $K_{TVrg}$ | 0.0000 | 0.0000 | 0.0000 | 0.0000 | 0.0000 |
| $K_{TVvo}$ | 0.0300 | 0.0300 | 0.0300 | 0.0300 | 0.0300 |
| $K_{TVvc}$ | 0.0400 | 0.0400 | 0.0400 | 0.0400 | 0.0400 |
| $K_{MVst}$ | 0.5000 | 0.5000 | 0.3000 | 0.6000 | 0.3000 |
| $K_{MVrg}$ | 0.0000 | 0.0000 | 0.0000 | 0.0000 | 0.0000 |
| $K_{MVvo}$ | 0.0300 | 0.0300 | 0.0300 | 0.0300 | 0.0300 |
| $K_{MVvc}$ | 0.0400 | 0.0400 | 0.0400 | 0.0400 | 0.0400 |
| $K_{AVst}$ | 0.5000 | 0.3000 | 0.3000 | 1.0000 | 0.3000 |
| $K_{AVrg}$ | 0.0000 | 0.0000 | 0.0000 | 0.0000 | 0.0000 |
| $K_{AVvo}$ | 0.1000 | 0.1000 | 0.1000 | 0.1000 | 0.1000 |
| $K_{AVvc}$ | 0.1000 | 0.1000 | 0.1000 | 0.1000 | 0.1000 |
| $K_{PVst}$ | 1.0000 | 1.0000 | 1.0000 | 0.8000 | 1.0000 |
| $K_{PVrg}$ | 0.0000 | 0.0000 | 0.0000 | 0.0000 | 0.0000 |
| $K_{PVvo}$ | 0.2000 | 0.2000 | 0.2000 | 0.2000 | 0.2000 |
| $K_{PVvc}$ | 0.2000 | 0.2000 | 0.2000 | 0.2000 | 0.2000 |
| $C_{Ao}$ | 0.0975 | 0.0882 | 0.0889 | 0.1240 | 0.1590 |
| $C_{Arc}$ | 0.0975 | 0.0882 | 0.0889 | 0.1240 | 0.1590 |
| $R_{AoA}$ | 0.4000 | 0.0100 | 0.1250 | 0.4910 | 0.5250 |
| $C_{UB}$ | 0.7590 | 0.5210 | 0.9510 | 0.8970 | 0.4490 |

| | | | | | |
|---|---|---|---|---|---|
| $L_{UBa}$ | 0.0007 | 0.0004 | 0.0007 | 0.0007 | 0.0007 |
| $R_{UBa}$ | 1.6200 | 2.1400 | 1.3700 | 1.4300 | 2.4000 |
| $R_{UBv}$ | 4.7200 | 6.2600 | 3.9900 | 4.1600 | 7.0000 |
| $C_{SVC}$ | 0.0741 | 0.0476 | 0.0838 | 0.0762 | 0.0337 |
| $R_{SVC}$ | 0.2160 | 0.3010 | 0.1970 | 0.2110 | 0.3900 |
| $C_{ThAo}$ | 0.0097 | 0.0122 | 0.0109 | 0.0100 | 0.0044 |
| $L_{ThAo}$ | 0.0024 | 0.0025 | 0.0023 | 0.0023 | 0.0023 |
| $R_{ThAo}$ | 0.5150 | 0.4330 | 0.4690 | 0.5040 | 0.9290 |
| $C_{AbAo}$ | 0.0205 | 0.0258 | 0.0232 | 0.0211 | 0.0093 |
| $L_{AbAo}$ | 0.0024 | 0.0025 | 0.0023 | 0.0023 | 0.0023 |
| $R_{AbAo}$ | 3.9200 | 3.3000 | 3.5800 | 3.8400 | 7.0800 |
| $C_{Lvr}$ | 0.2550 | 0.3220 | 0.2890 | 0.2630 | 0.1160 |
| $R_{Lvra}$ | 61.2000 | 51.5000 | 55.8000 | 59.9000 | 110.0000 |
| $R_{Lvrv}$ | 0.3830 | 0.3220 | 0.3490 | 0.3750 | 0.6920 |
| $C_{Kd}$ | 0.1060 | 0.1340 | 0.1200 | 0.1090 | 0.0483 |
| $R_{Kda}$ | 43.2000 | 36.3000 | 39.4000 | 42.3000 | 77.9000 |
| $R_{Kdv}$ | 4.0400 | 3.4000 | 3.6800 | 3.9600 | 7.2900 |
| $C_{I}$ | 0.0636 | 0.0801 | 0.0719 | 0.0654 | 0.0289 |
| $R_{Ia}$ | 94.6000 | 79.6000 | 86.3000 | 92.7000 | 171.0000 |
| $R_{Iv}$ | 1.7200 | 1.4500 | 1.5700 | 1.6800 | 3.1000 |
| $C_{Lega}$ | 0.0067 | 0.0076 | 0.0065 | 0.0055 | 0.0020 |
| $L_{Lega}$ | 0.0024 | 0.0025 | 0.0023 | 0.0023 | 0.0023 |

| | | | | | |
|---|---|---|---|---|---|
| $R_{Lega}$ | 14.4000 | 13.1000 | 14.8000 | 16.6000 | 35.5000 |
| $R_{Legc}$ | 33.8000 | 30.8000 | 34.8000 | 39.1000 | 83.6000 |
| $C_{Legv}$ | 0.1370 | 0.1550 | 0.1320 | 0.1130 | 0.0410 |
| $R_{Legv}$ | 7.9100 | 7.2100 | 8.1500 | 9.1500 | 19.6000 |
| $C_{AbIV}$ | 0.1060 | 0.1330 | 0.1190 | 0.1090 | 0.0480 |
| $R_{AbIV}$ | 0.1080 | 0.0907 | 0.0983 | 0.1060 | 0.1950 |
| $C_{ThIV}$ | 0.3160 | 0.3980 | 0.3580 | 0.3250 | 0.1440 |
| $R_{ThIV}$ | 0.0840 | 0.0707 | 0.0766 | 0.0823 | 0.1520 |
| $R_{PDA}$ | 117.0000 | 3.1700 | 0.4000 | $10^5$ | 1.5000 |
| $C_{PA}$ | 0.1900 | 0.1710 | 0.0884 | 0.1200 | 0.0884 |
| $C_{PA2}$ | 0.1900 | 0.1710 | 0.0884 | 0.1200 | 0.0884 |
| $R_T$ | 0.0100 | 0.0100 | 0.0020 | 0.0020 | 0.0020 |
| $RCRr_{C1}$ | 0.2140 | 0.1290 | 0.0717 | 0.1520 | 0.0816 |
| $RCRr_{C2}$ | 0.0439 | 0.0265 | 0.0147 | 0.0313 | 0.0168 |
| $RCRr_{C3}$ | 1.9900 | 1.2000 | 0.6670 | 1.4200 | 0.7590 |
| $RCRr_{Rd1}$ | 4.2400 | 6.1800 | 9.6200 | 4.3300 | 8.7300 |
| $RCRr_{Rd2}$ | 17.9000 | 26.1000 | 40.7000 | 18.3000 | 36.9000 |
| $RCRr_{Rd3}$ | 0.5720 | 0.8340 | 1.3000 | 0.5840 | 1.1800 |
| $RCRr_{Rp1}$ | 0.8690 | 1.2700 | 1.9700 | 0.8860 | 1.7900 |
| $RCRr_{Rp2}$ | 2.2200 | 3.2300 | 5.0300 | 2.2600 | 4.4600 |
| $RCRr_{Rp3}$ | 0.2820 | 0.4110 | 0.6390 | 0.2880 | 0.5800 |
| $RCRl_{C1}$ | 0.0333 | 0.0202 | 0.0112 | 0.0237 | 0.0127 |

| | | | | | |
|---|---|---|---|---|---|
| $RCRl_{C2}$ | 0.0821 | 0.0497 | 0.0276 | 0.0556 | 0.0314 |
| $RCRl_{C3}$ | 1.0400 | 0.6270 | 0.3480 | 0.7390 | 0.3950 |
| $RCRl_{Rd1}$ | 25.8000 | 37.5000 | 58.4000 | 26.3000 | 53.0000 |
| $RCRl_{Rd2}$ | 10.9000 | 15.9000 | 24.8000 | 11.2000 | 22.5000 |
| $RCRl_{Rd3}$ | 1.3700 | 2.0000 | 3.1100 | 1.4000 | 2.8300 |
| $RCRl_{Rp1}$ | 0.5260 | 0.7660 | 1.1900 | 0.5360 | 1.0800 |
| $RCRl_{Rp2}$ | 0.2230 | 0.3250 | 0.5060 | 0.2280 | 0.4590 |
| $RCRl_{Rp3}$ | 0.0572 | 0.0834 | 0.1300 | 0.0584 | 0.1180 |

**Units:** ***T*** (Cardiac cycle duration, *s*), ***c*** (Elastance factor of the cardiac chambers, *mmHg*), ***d*** (Diastolic exponential factor of the cardiac chambers, $cm^{-3}$), $\boldsymbol{V_0}$ (Unstressed volume, $cm^3$), ***a*** (Quadratic elastance factor for the ventricles, $mmHg.cm^{-6}$), ***b*** (Linear elastance factor for the ventricles, $mmHg.cm^{-3}$), ***R*** (Vascular resistance, $mmHg.s.cm^{-3}$), ***L*** (Vascular inductance/inertia, $mmHg.s^2.cm^{-3}$), $\boldsymbol{RCR_C}$ (Compliance of the pulmonary RCR block, $cm^3.mmHg^{-1}$), $\boldsymbol{RCR_{Rd}}$ (Distal resistance of the pulmonary RCR block, $mmHg.s.cm^{-3}$), $\boldsymbol{RCR_{Rp}}$ (Proximal resistance of the pulmonary RCR block, $mmHg.s.cm^{-3}$); †patients underwent pulmonary artery banding; †patients underwent pulmonary artery banding. †data collected based on echocardiography alone;

| | | | | | |
|---|---|---|---|---|---|
| (grey) | Fixed clinical inputs | (orange) | Automatic tuning framework | (blue) | Parameters from scaling |

**SUPPLEMENTARY TABLE ST6. Hemodynamic quantities predicted by the preoperative LPN model for the patients in Group I. The error in LPN prediction, measured as a (%) of the clinical data, is shown in parentheses.**

| **Patient** | **A*** | **B*** | **C** | **D** | **E** |
|---|---|---|---|---|---|
| **$AOP_{min}$** | 42.66 **(4.1%)** | 35.53 **(4.0%)** | 32.69 **(3.9%)** | 26.25 **(6.3%)** | 30.02 **(9.0%)** |
| **$AOP_{max}$** | 63.28 **(2.7%)** | 95.04 **(1.1%)** | 90.96 **(5.3%)** | 49.78 **(4.3%)** | 64.60 **(3.6%)** |
| **$AOP_{mean}$** | 52.75 **(7.7%)** | 55.69 **(7.2%)** | 50.96 **(3.9%)** | 34.80 **(3.3%)** | 43.97 **(0.1%)** |
| **$PAP_{min}$** | 36.24 **(6.6%)** | 34.88 **(3.1%)** | 31.99 **(3.2%)** | 27.12 **(0.4%)** | 27.68 **(2.5%)** |
| **$PAP_{max}$** | 59.46 **(1.0%)** | 103.43 **(4.2%)** | 53.50 **(0.9%)** | 43.78 **(9.5%)** | 52.69 **(7.5%)** |
| **$PAP_{mean}$** | 45.76 **(4.0%)** | 55.88 **(8.4%)** | 40.62 **(0.9%)** | 33.59 **(4.0%)** | 34.91 **(8.4%)** |
| **$LAP_{mean}$** | 5.33 **(6.6%)** | 4.97 **(0.6%)** | 16.02 **(30.4%)** | 12.39 **(11.5%)** | 11.92 **(8.4%)** |
| **Qp/Qs** | 0.95 **(3.0%)** | 0.97 **(9.4%)** | 1.87 **(3.1%)** | 2.91 **(2.0%)** | 2.01 **(0.5%)** |
| **PVRi** | 2.54 **(1.6%)** | 3.53 **(2.6%)** | 4.12 **(0.5%)** | 3.06 **(1.3%)** | 2.60 **(8.3%)** |
| **SVRi** | 11.10 **(4.3%)** | 13.24 **(4.2%)** | 12.64 **(0.3%)** | 9.98 **(0.8%)** | 5.61 **(4.9%)** |
| **$MVPG_{max}$** | 5.77 **(3.8%)** | 8.23 **(8.5%)** | 14.83 **(12.8%)** | 8.41 **(5.1%)** | 9.70 **(11.8%)** |
| **$AVPG_{max}$** | - | - | - | 11.20 **(1.3%)** | 26.14 **(0.5%)** |
| **$PVPG_{max}$** | - | 7.10 **(11.3%)** | - | - | - |
| **$ASDPG_{mean}$** | 1.00 **(0.0%)** | 1.09 **(9.0%)** | 10.00 **(0.0%)** | 5.88 **(2.0%)** | 2.64 **(5.6%)** |
| **$PDAPG_{mean}$** | **6.99 (39.8%)** | - | 10.34 **(13.8%)** | 1.20 **(20.0%)** | 9.06 **(0.7%)** |

*patients underwent pulmonary artery banding; All pressures and pressure gradients are in *mmHg*; PVRi/SVRi: $WU{\cdot}m^2$.

**SUPPLEMENTARY TABLE ST7. Hemodynamic quantities predicted by the preoperative LPN model for the patients in Group II. The error in LPN prediction, measured as a (%) of the clinical data, is shown in parentheses.**

| **Patient** | **F** | **G** | **H†** | **I†** | **J†** |
|---|---|---|---|---|---|
| **$AOP_{min}$** | 41.72 **(6.9%)** | **39.86 (11.4%)** | 38.60 **(5.9%)** | 39.70 **(1.8%)** | 42.92 **(10.6%)** |
| **$AOP_{max}$** | 71.60 **(4.5%)** | 69.54 **(5.4%)** | 61.81 **(9.1%)** | 56.02 **(6.6%)** | 68.30 **(1.9%)** |
| **$AOP_{mean}$** | 53.81 **(0.4%)** | 53.10 **(5.2%)** | 48.39 **(3.2%)** | 46.14 **(0.3%)** | 52.56 **(4.4%)** |
| **$PAP_{min}$** | 15.58 **(2.6%)** | 22.51 **(2.1%)** | 31.15 **(2.7%)** | 15.89 **(5.9%)** | 28.84 **(0.6%)** |
| **$PAP_{max}$** | 29.91 **(0.3%)** | 32.26 **(7.8%)** | 50.97 **(8.5%)** | 25.31 **(6.3%)** | 34.35 **(7.2%)** |
| **$PAP_{mean}$** | 19.93 **(9.4%)** | 25.49 **(8.9%)** | 38.38 **(1.6%)** | 20.07 **(5.6%)** | 31.28 **(5.2%)** |
| **$LAP_{mean}$** | 3.73 **(25.4%)** | 8.72 **(20.7%)** | 13.42 **(3.2%)** | 4.74 **(5.2%)** | 15.59 **(19.9%)** |
| **Qp/Qs** | 3.74 **(1.1%)** | 3.18 **(6.5%)** | 3.33 **(3.1%)** | 3.81 **(7.9%)** | 3.06 **(6.1%)** |
| **PVRi** | 1.84 **(3.2%)** | 2.68 **(8.5%)** | 3.65 **(4.6%)** | 1.57 **(7.1%)** | 2.75 **(11.9%)** |
| **SVRi** | 19.35 **(1.3%)** | 22.23 **(6.1%)** | 29.74 **(6.5%)** | **14.96 (16.4%)** | 22.39 **(2.1%)** |
| **SV (mL)** | - | - | **4.30 (34.4%)** | 3.40 **(6.3%)** | **3.91 (39.6%)** |
| **EF (%)** | - | 57.90 **(0.2%)** | 63.40 **(0.9%)** | 50.90 **(1.8%)** | 64.70 **(17.6%)** |
| **$MVPG_{max}$** | 7.29 **(8.9%)** | - | 7.17 **(19.5%)** | 8.32 **(2.1%)** | 11.31 **(13.1%)** |
| **$AVPG_{max}$** | 14.31 **(30.1%)** | - | **20.00 (53.85%)** | 1.82 **(9.0%)** | **43.00 (30.3%)** |
| **$PVPG_{max}$** | - | 5.01 **(16.5%)** | 8.32 **(4.0%)** | 14.10 **(17.5%)** | 1.80 **(10.0%)** |
| **$VSDPG_{mean}$** | - | - | **30.74 (33.7%)** | 20.64 **(6.18%)** | - |
| **$ASDPG_{mean}$** | **1.00 (50.0%)** | 7.04 **(12.00%)** | 10.00 **(11.1%)** | 0.82 **(18.00%)** | 13.21 **(10.0%)** |
| **$PDAPG_{mean}$** | 34.27 **(7.1%)** | **27.61 (25.5%)** | 10.02 **(8.9%)** | - | 21.28 **(3.3%)** |

†data collected based on echocardiography alone; All pressures and pressure gradients are in *mmHg*; PVRi/SVRi: $WU{\cdot}m^2$.

**SUPPLEMENTARY TABLE ST8. Predicted hemodynamic variables following virtual BiVR and S1P surgical procedures for the patients in Group I.**

| Patient / Hemodynamic Variable | Surgery | A | B | C | D | E |
|---|---|---|---|---|---|---|
| $PAP_{max}$(mmHg) | BiVR | 28.00 | 48.00 | 48.00 | 38.00 | 65.00 |
| | S1P* | 14.00 | 18.00 | 22.50 | 19.00 | 19.00 |
| $PAP_{mean}$ (mmHg) | BiVR | 24.00 | 41.00 | 39.00 | 32.00 | 54.00 |
| | S1P* | 13.00 | 17.00 | 21.50 | 18.00 | 18.00 |
| $AOP_{max}$ (mmHg) | BiVR | 43.00 | 42.00 | 72.00 | 59.00 | 49.00 |
| | S1P* | 91.00 | 99.00 | 82.00 | 67.00 | 82.00 |
| $AOP_{mean}$ (mmHg) | BiVR | 38.00 | 35.00 | 48.00 | 43.03 | 39.00 |
| | S1P* | 49.00 | 58.00 | 52.00 | 43.00 | 50.00 |
| $LAP_{mean}$ (mmHg) | BiVR | 17.00 | 32.00 | 21.00 | 19.00 | 38.00 |
| | S1P* | 5.00 | 4.50 | 7.00 | 7.00 | 7.50 |
| SVEDP (mmHg) | BiVR | 19.00 | 30.00 | 14.00 | 12.00 | 34.00 |
| | S1P* | 5.00 | 3.00 | 5.00 | 7.00 | 4.00 |
| SVCP (mmHg) | BiVR | 2.49 | 2.80 | 2.92 | 3.58 | 4.02 |
| | S1P* | 6.23 | 7.13 | 5.67 | 7.65 | 8.27 |
| Qp/Qs | BiVR | 0.99 | 1.01 | 1.00 | 1.00 | 1.00 |
| | S1P* | 0.84 | 0.92 | 0.85 | 0.80 | 0.53 |
| Qs (L/min) | BiVR | 0.93 | 0.66 | 0.98 | 0.89 | 1.26 |
| | S1P* | 1.07 | 1.22 | 0.96 | 0.75 | 1.59 |
| iQs (L/min/$m^2$) | BiVR | 3.20 | 2.53 | 3.62 | 4.23 | 6.00 |
| | S1P* | 3.69 | 4.69 | 3.55 | 3.57 | 7.57 |
| CO (L/min) | BiVR | 0.93 | 0.66 | 0.98 | 0.89 | 1.26 |
| | S1P* | 1.96 | 2.02 | 1.78 | 1.35 | 2.44 |
| CI (L/min/$m^2$) | BiVR | 3.20 | 2.53 | 3.62 | 4.23 | 6.00 |
| | S1P* | 6.75 | 7.77 | 6.59 | 6.43 | 11.61 |
| $S_{art}$ (%) | BiVR | 98.00 | 98.00 | 98.00 | 98.00 | 98.00 |

| | | | | | | |
|---|---|---|---|---|---|---|
| | S1P* | 72.00 | 77.00 | 72.00 | 75.00 | 78.00 |
| **$S_{ven}$ (%)** | BiVR | 73.00 | 66.00 | 76.00 | 79.00 | 83.00 |
| | S1P* | 50.00 | 57.00 | 49.00 | 53.00 | 68.00 |
| **$O_2D$ ($mlO_2/min/m^2$)** | BiVR | 671.00 | 530.00 | 759.00 | 886.00 | 1238.00 |
| | S1P* | 558.00 | 665.00 | 545.00 | 574.00 | 1272.00 |

*indicates clinically performed procedure for the particular group.

**SUPPLEMENTARY TABLE ST9. Predicted hemodynamic variables following virtual BiVR and S1P surgical procedures for the patients in Group II.**

| Patient / Hemodynamic Variable | Surgery | F | G | H | I | J |
|---|---|---|---|---|---|---|
| **$PAP_{max}$ (mmHg)** | BiVR* | 18.00 | 21.00 | 26.00 | 17.00 | 21.00 |
| | S1P | 13.00 | 16.50 | 21.00 | 12.00 | 20.00 |
| **$PAP_{mean}$ (mmHg)** | BiVR* | 12.00 | 14.00 | 20.00 | 14.00 | 17.00 |
| | S1P | 12.00 | 16.00 | 19.00 | 11.00 | 19.00 |
| **$AOP_{max}$ (mmHg)** | BiVR* | 98.00 | 98.50 | 81.00 | 72.00 | 92.00 |
| | S1P | 126.00 | 106.00 | 98.00 | 84.00 | 106.00 |
| **$AOP_{mean}$ (mmHg)** | BiVR* | 82.00 | 86.00 | 66.00 | 62.00 | 75.00 |
| | S1P | 75.00 | 74.00 | 64.00 | 53.00 | 71.00 |
| **$LAP_{mean}$ (mmHg)** | BiVR* | 3.64 | 3.40 | 10.00 | 7.53 | 6.23 |
| | S1P | 3.00 | 3.00 | 3.00 | 4.00 | 3.00 |
| **SVEDP (mmHg)** | BiVR* | 4.00 | 2.00 | 5.00 | 4.00 | 3.00 |
| | S1P | 2.36 | 2.50 | 3.00 | 3.00 | 3.00 |
| **SVCP (mmHg)** | BiVR* | 2.50 | 3.02 | 4.38 | 3.50 | 4.06 |
| | S1P | 5.07 | 4.57 | 7.19 | 5.83 | 5.56 |
| **Qp/Qs** | BiVR* | 0.99 | 0.99 | 1.00 | 1.00 | 1.01 |
| | S1P | 1.33 | 1.50 | 2.10 | 1.44 | 2.01 |
| **Qs (L/min)** | BiVR* | 1.00 | 0.91 | 0.57 | 0.82 | 0.64 |
| | S1P | 0.93 | 0.79 | 0.48 | 0.69 | 0.54 |
| **iQs (L/min/$m^2$)** | BiVR* | 4.00 | 3.96 | 2.38 | 3.91 | 3.39 |
| | S1P | 3.72 | 3.43 | 2.00 | 3.29 | 3.00 |
| **CO (L/min)** | BiVR* | 1.00 | 0.91 | 0.57 | 0.82 | 0.64 |
| | S1P | 2.16 | 1.97 | 1.50 | 1.68 | 1.64 |
| **CI (L/min/$m^2$)** | BiVR* | 4.00 | 3.96 | 2.38 | 3.91 | 3.39 |
| | S1P | 8.64 | 8.57 | 6.25 | 8.00 | 9.11 |
| | BiVR* | 98.00 | 98.00 | 98.00 | 98.00 | 98.00 |

| | | | | | | |
|---|---|---|---|---|---|---|
| **$S_{art}$ (%)** | S1P | 82.00 | 83.00 | 80.00 | 81.00 | 85.00 |
| **$S_{ven}$ (%)** | BiVR* | 78.00 | 78.00 | 64.00 | 78.00 | 76.00 |
| | S1P | 60.00 | 59.00 | 40.00 | 57.00 | 59.00 |
| **$O_2D$ ($mlO_2/min/m^2$)** | BiVR* | 833.00 | 826.00 | 494.00 | 819.00 | 744.00 |
| | S1P | 647.00 | 602.00 | 358.00 | 566.00 | 549.00 |

*indicates clinically performed procedure for the particular group.

### SUPPLEMENTARY FIGURES

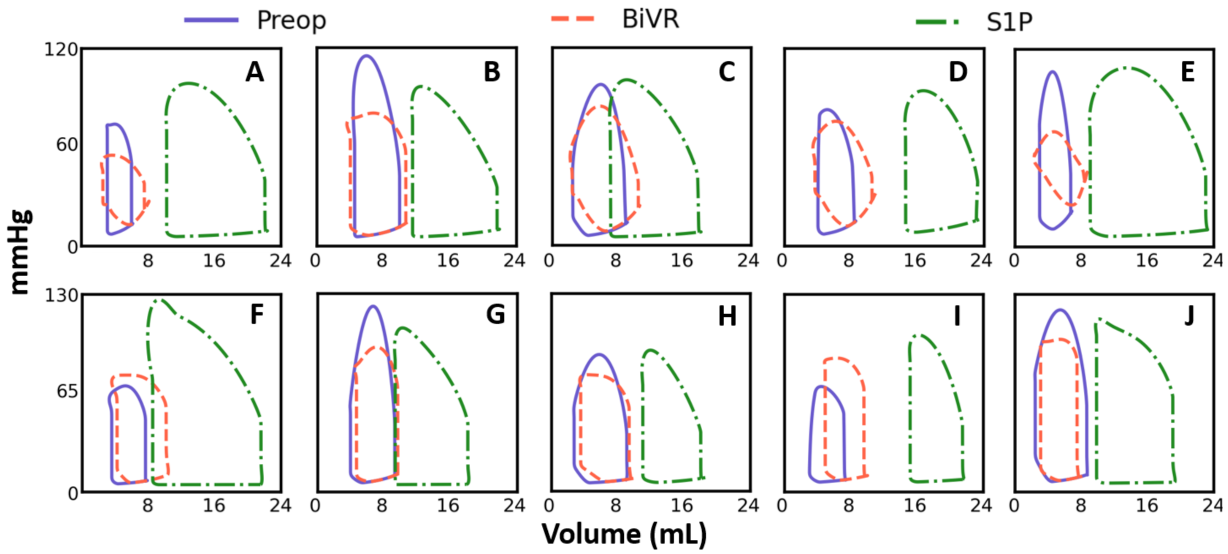


**Supplementary Figure SF1.** Comparison of the LPN model-predicted pressure-volume loops of the systemic ventricle for all patients in the study cohort between preoperative (**preop**) and virtual surgeries (**BiVR**: biventricular repair; **S1P**: stage-1 palliation). (**top**) A-E are Group I patients who have clinically undergone S1P. (**bottom**) F-J are Group II patients who underwent BiVR clinically.

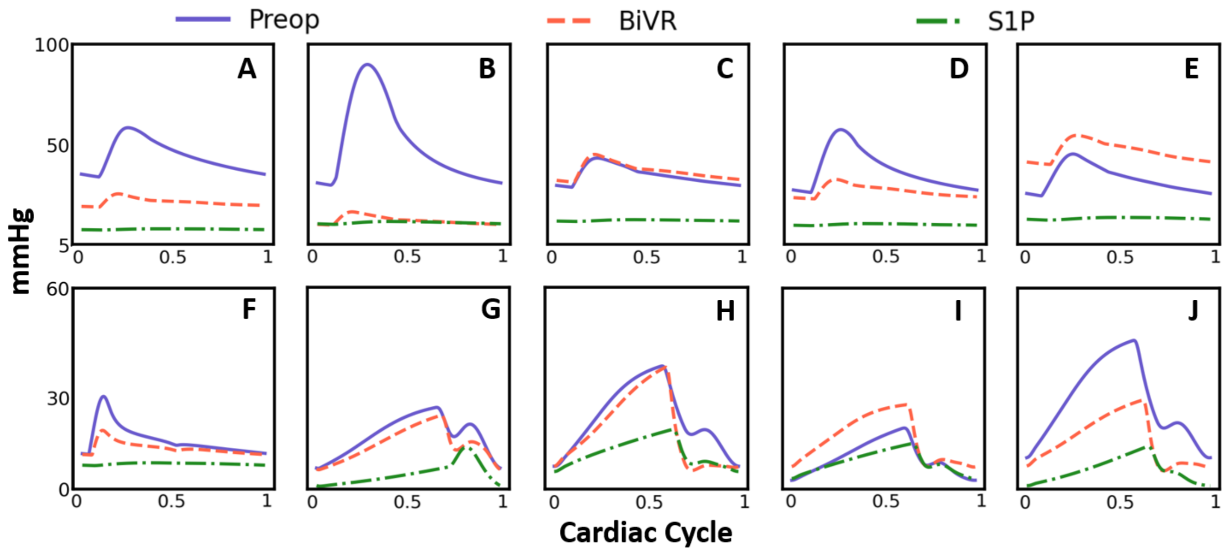


**Supplementary Figure SF2.** Comparison of predicted pulmonary artery pressure waveforms during the cardiac cycle for all patients in the study cohort between preoperative (**preop**) and virtual surgeries (**BiVR**: biventricular repair; **S1P**: stage-1 palliation). (**top**) A-E are Group I patients who have clinically undergone S1P. (**bottom**) F-J are Group II patients who underwent BiVR clinically.

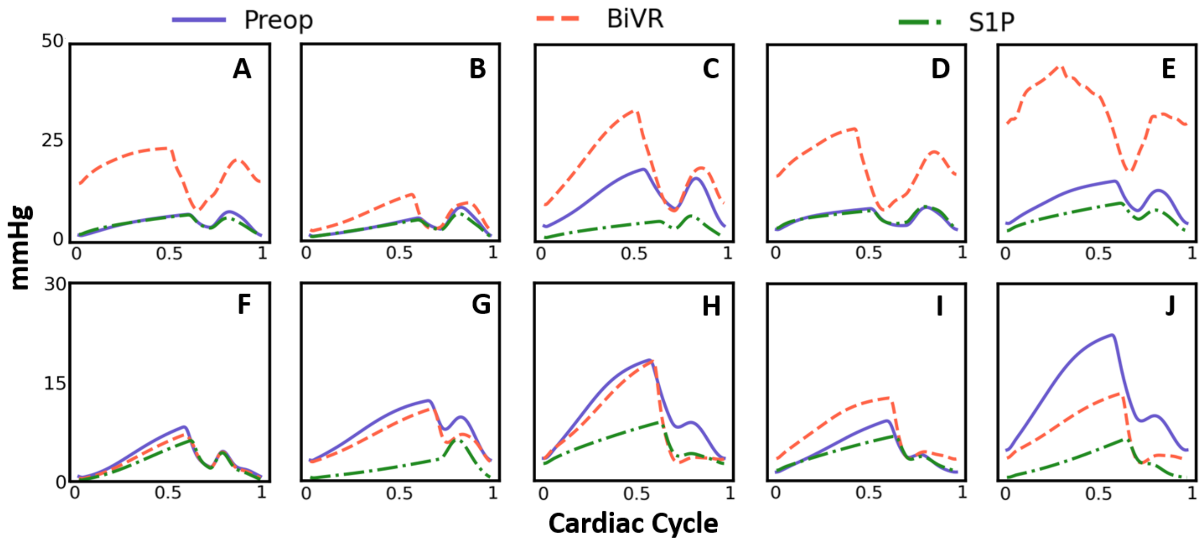


**Supplementary Figure SF3.** Comparison of predicted left atrial pressure waveforms during the cardiac cycle for all patients in the study cohort between preoperative (**preop**) and virtual surgeries (**BiVR**: biventricular repair; **S1P**: stage-1 palliation). (**top**) A-E are Group I patients who have clinically undergone S1P. (**bottom**) F-J are Group II patients who underwent BiVR clinically
